# Causal effect of video gaming on mental well-being in Post-COVID Japan

**DOI:** 10.64898/2026.08.12.26360262

**Authors:** Hiroyuki Egami, Md. Shafiur Rahman, Chihiro Egami, Tsuyoshi Yamamoto, Takahisa Wakabayashi, Shunsuke Horii, Andrew K. Przybylski

**Author notes:** Corresponding author Hiroyuki Egami, PhD, University of Oxford, Oxford, The United Kingdom.

## Abstract

**IMPORTANCE:** With a growing global user base of 3.5 billion and users spending nearly as much time gaming as on social media, video gaming’s effects on mental well-being have attracted scholarly and public interest. Despite the WHO’s inclusion of gaming disorder in ICD-11 and government-mandated restrictions in multiple countries, the causal evidence supporting such policies remains limited.

**OBJECTIVE:** To investigate the causal effect of video gaming on mental well-being in the post-COVID period.

**DESIGN:** A natural experiment of game console lottery was used to identify the causal effect of video gaming on mental well-being. The intention-to-treat effect was estimated using multivariate regression and propensity score matching. Causal effects of game engagement were estimated using the instrumental variable method (two-stage least squares) and a causal machine learning algorithm—instrumental forest.

**SETTING:** Online suarveys were conducted between September 2022 and March 2023, covering all 47 prefectures in Japan.

**PARTICIPANTS:** A total of 71,435 participants aged 10-69 answered the surveys. 6,911 individuals participated in the natural experiment.

**EXPOSURES:** Video game engagement, including video game console ownership, use of the console in the last 30 days, and video gaming duration.

**MAIN OUTCOMES AND MEASURES:** Psychological distress and life satisfaction.

**RESULTS:** The intention-to-treat effects of winning game console lotteries on mental well-being were positive (0.1 SD). Game console ownership improved mental well-being by 0.1–0.2 SD, and past-month play improved it by 0.2–0.3 SD. An extra hour of daily video game play led to 0.3-0.5 SD improvements in mental well-being.

**CONCLUSIONS AND RELEVANCE:** This study found that video gaming had a positive effect on mental well-being in the post-COVID period. The consistency of the effect size with that of a related COVID-period study adds robustness to our findings. Our findings add to the growing evidence that digital media screen time has diverse effects on well-being and support public health policies that recognize the potential mental well-being benefits of appropriate levels of video gaming.

**Key points:** *Questions:* Is there a causal relationship between video gaming and mental well-being among the Japanese population in the post-COVID period?

*Findings:* A natural experiment in post-COVID Japan demonstrated that video game engagement reduced psychological distress and improved life satisfaction, with effect sizes broadly consistent with those observed during the COVID-19 period.

*Meaning:* Video gaming appears to benefit mental well-being not only during crisis periods but also in non-crisis contexts. This underscores the importance of accumulating causal evidence and suggests that moderate video game engagement may contribute positively to mental well-being.

## Introduction

Digital media may profoundly affect human behavior and well-being, with video gaming emerging as a particularly significant medium. Gaming’s societal impact extends far beyond mere entertainment. Video games have reached over 3.5 billion players globally.^1^ Users spend nearly as much time gaming as they do on social media.^2^ Alongside gaming’s rising prominence in daily life, public concerns about its potentially harmful effects on well-being have grown, prompting significant institutional responses. For instance, the World Health Organization (WHO) included Gaming Disorder in the International Classification of Diseases (ICD-11) and advised users to be aware of how much time they spend gaming.^3^ Furthermore, several government authorities, including the Chinese government^4^ and local governments in Japan,^5^ have attempted to limit gaming time.

Numerous empirical studies have studied the relationship between video gaming and mental well-being, yet the implications have been mixed. Some have suggested a positive link,^6–15^ others a statistically insignificant relationship,^16–23^ while still others have suggested a negative link.^24–29^ This body of work includes both observational studies and laboratory experiments, each facing different methodological limitations. Observational studies are predominantly associational and, therefore, have limited capacity to establish causality between gaming and well-being. Meanwhile, laboratory experiments have limited capacity to assess the effects of gaming as an integrated lifestyle rather than an isolated activity, because it is difficult to replicate real-world gaming contexts in a lab.

Our previous work^30^ employed a natural experiment approach to analyze observational data covering the age range of 10-69 in Japan, establishing a new benchmark for evidence of the mental effects of digital media. A natural experiment is a methodologically promising alternative because it can establish causality from real-world data. By examining the effect of owning a game console, our study demonstrated that video gaming has positive effects on mental well-being. This finding is supported by the literature, which has demonstrated several possible positive pathways linking gaming to mental well-being, including the promotion of social capital, exergaming, and stress coping.^31^ However, our previous study^30^ analyzed data collected during the COVID-19 pandemic period, which limits the generalizability of its findings. Although the potential loss of outdoor activity time due to video gaming could be a negative pathway of gaming’s effect under ordinary conditions, people had fewer opportunities to play outside during the COVID-19 pandemic. As such, a negative pathway might have worked minimally in the context; the positive effect of gaming might have been overestimated.

Further empirical studies investigating the causal relationship between video gaming and mental well-being across different social contexts are essential to strengthen the evidence base for clinical applications and regulatory discussions. However, apart from our previous study,^30^ no research has explicitly examined the causal relationship between gaming and mental well-being using causal inference methods with real-world data. Therefore, to address this gap, this study conducted another natural experiment in a typical social context, utilizing post-COVID observational data from Japan, to investigate the causal effects of video gaming on mental health outcomes.

## Methods

### Study design

We conducted a natural experimental study, using a game console lottery that ran from 2020 to 2023 in Japan, to identify the causal effect of video gaming on mental well-being. During this period, the supply of PlayStation5 (PS5), a major gaming console, was severely constrained, making it impossible for all interested consumers to purchase one. Japanese retailers used lotteries to assign PS5s to consumers, inadvertently creating a plausibly random variation in ownership. Winning the lottery was a primary determinant of PS5 ownership at that time. We leveraged this situation as a natural experiment to draw causal inferences.

### Study participants and sources of data

The study participants were people aged 10–69 from all 47 prefectures in Japan (n = 71,435). In collaboration with the gaming research firm gameage R&I (GRI) and the survey firm Cross Marketing, we conducted three rounds of omnibus online surveys between September 2022 and March 2023. A stratified random sampling technique was used to recruit participants from the partner companies’ pool of 316,985 respondents (strata: age group, gender and video gaming preferences). The response rate was 46.6% (Supplementary Table 1).

We collected information on participants’ lottery participation, video game ownership, gaming duration, gaming preferences, sociodemographic characteristics, mental health, and life satisfaction. After excluding people who did not participate in a PS5 lottery, the analysis sample for our causal inference consisted of 6,911 responses from individuals aged 10–69 years (Supplementary Fig 1). Our dataset is a blend of panel and repeated cross-sectional observations. This data structure facilitates the validation of natural experiments. Further details on lottery context and data collection are provided in eMethods.

### Measurements

#### Exposures

The main exposure was video game engagement (Supplementary Table 2). Respondents were asked whether their households owned PS5 and whether they had played one in the past 30 days. Additionally, respondents reported the time spent playing video games on weekdays and weekends over the past 30 days (asked separately). We collected data separately on time spent playing (1) video games on a TV or computer (encompassing PS5) and (2) smartphone games.

#### Outcomes

The primary outcomes were two aspects of well-being: mental health and life satisfaction. Participants’ mental health status was assessed using the K6 scale^32,33^, which features six items that assess nonspecific psychological distress over the past 30 days, with a total score ranging from 0 (minimum distress) to 24 (maximum distress). Given its reliability, the K6 is employed in population-based health surveys in Japan as a screening tool for mental distress.^33^ Life satisfaction was measured using the Japanese version of the Satisfaction with Life Scale (SWLS).^34,35^ This 5-item scale is scored between 1 (completely disagree) and 7 (completely agree), with a total score ranging from 5 (lowest) to 35 (highest). The SWLS is a reliable and validated instrument for assessing overall life satisfaction in Japan.

#### Covariates

Following our previous study,^30^ we employed a set of covariates (Supplementary Table 3) including those of respondents: age, gender, employment status, residential prefecture, number of times that respondent households entered game console lotteries, and video gaming preference. Additionally, we utilized the characteristics of either respondents or caregivers as covariates: marital status, having children (yes, no) and occupation.

### Statistical analysis

We used four methods: multivariate regression, propensity score matching (PSM) approach, instrumental variable (IV) method, and a causal machine learning algorithm called causal forest (or generalized random forests, GRFs).^36,37^ These methods are consistent with our previous study.^30^

#### Multivariate regression and PSM

We employed multivariate regression and PSM to estimate the causal effect of winning game console lotteries. Namely, we conducted an intention-to-treat (ITT) analysis (i.e., a reduced form analysis), following previous lottery-based natural experiment studies.^38,39^ A treatment assignment variable, a binary variable equal to 1 when a participant won a PS5 lottery, was employed. We assumed unconfoundedness for a PS5 lottery, controlling for the number of lottery entries. See eMethods for details. We further examined whether purchasing a new video game console would increase video game play duration. Time spent on smartphone gaming was also analyzed as an additional outcome.

#### Assessing natural experiment validity

Among the underlying assumptions for establishing causality, notably, the unconfoundedness (or conditional exchangeability) is ensured by the natural experimental design. We assessed the unconfoundedness of the PS5 lotteries and validated our study design, using two methods. First, we used standardized differences and assessed the balance of baseline characteristics. Second, we used pseudo-outcome tests, utilizing pre-lottery well-being values as pseudo-outcomes. Further details are provided in eMethods.

#### IV approach

We used the two-stage least squares (2SLS) estimation to estimate the causal effect of owning PS5—Local Average Treatment Effect (LATE). We treated lottery wins as an excluded instrument to address the unmeasured confounding bias. As exposure variables, we examined PS5 ownership, playing PS5 last month, and gameplay duration. The lottery-winning instrument plausibly satisfies the exclusion restriction for PS5 ownership, though reservations remain for other game-play-related variables. See eMethods for details.

#### Machine learning: Instrumental forest

To assess the robustness of IV regression estimates, we estimated the causal effect of game engagement (namely, LATE) using instrumental forest algorithm, which incorporates IV approach to generalized random forests (GRF).^36,37^ GRF provides various non-parametric estimation methods that rely less on the assumption of a linear relationship between independent and dependent variables. Given the trained instrumental forest, GRF can also estimate LATE (or, more specifically, Average Conditional Local Average Treatment Effect, ACLATE) by averaging doubly robust scores for each training sample, which is constructed via augmented Inverse-Propensity Weighting (AIPW). More details on instrumental forests are shown in eMethods and elsewhere.^40^

#### Sensitivity analysis

We conducted a sensitivity analysis by excluding data collected during the September 2022 survey round. While we consider the three survey rounds (conducted in September and December 2022, and March 2023) as a post-COVID period (see our survey schedule in Supplementary Fig 9), it is unclear when the COVID period ended. Therefore, to assess the sensitivity of the results, we excluded the September 2022 survey round and used the remaining two survey rounds.

## Results

### Participant characteristics

A total of 71,435 respondents aged 10-69 years were included in this study (Table 1). One-fourth of the respondents were aged 10-24 years. In addition, 22% were students, 10% were unemployed, and 40% were full-time employees. Of the 71,435 survey respondents, 6,911 entered the lottery. One-third of the lottery participants (32%) were hardcore gamers, and 31% were core gamers. Video gaming preference was closely related to time spent playing games, with hardcore gamers playing for more than 1 h 20 min per day (Supplementary Table 4).

**Table 1:** Background characteristics of study participants.

|  | Lottery participant status |  | Total |
| --- | --- | --- | --- |
|  | PS5 lottery participant | Non-participant |  |
| Total | 6,911 (100.0) | 64,524 (100.0) | 71,435 (100.0) |
| Age |  |  |  |
| 10-24 years (n, %) | 1,696 (24.5) | 15,908 (24.7) | 17,604 (24.6) |
| 25-44 years | 2,609 (37.8) | 22,422 (34.7) | 25,031 (35.0) |
| 45-69 years | 2,606 (37.7) | 26,194 (40.6) | 28,800 (40.3) |
| Male | 4,268 (61.8) | 33,141 (51.4) | 37,409 (52.4) |
| Have children (yes) | 3,721 (53.9) | 32,391 (50.2) | 36,112 (50.6) |
| Marital status |  |  |  |
| Married | 4,255 (61.6) | 36,650 (56.8) | 40,905 (57.3) |
| Divorced/separated | 417 (6.0) | 4,408 (6.8) | 4,825 (6.8) |
| Not married | 2,231 (32.3) | 23,424 (36.3) | 25,655 (35.9) |
| Occupation |  |  |  |
| Student | 1,522 (22.0) | 14,022 (21.7) | 15,544 (21.8) |
| Stay-at-home wife/husband | 534 (7.7) | 6,304 (9.8) | 6,838 (9.6) |
| Full-time employee | 3,311 (47.9) | 25,113 (38.9) | 28,424 (39.8) |
| Part-time employee | 606 (8.8) | 8,045 (12.5) | 8,651 (12.1) |
| Self employed/others | 483 (7.0) | 4,199 (6.5) | 4,682 (6.6) |
| Unemployed/not a student | 455 (6.6) | 6,841 (10.6) | 7,296 (10.2) |
| Gaming preference |  |  |  |
| Hardcore gamer | 2,237 (32.4) | 8,959 (13.9) | 11,196 (15.7) |
| Core gamer | 2,121 (30.7) | 15,133 (23.5) | 17,254 (24.2) |
| Middle-core gamer | 1,441 (20.9) | 17,523 (27.2) | 18,964 (26.5) |
| Casual gamer | 838 (12.1) | 16,741 (25.9) | 17,579 (24.6) |
| Non-gamer | 274 (4.0) | 6,168 (9.6) | 6,442 (9.0) |
| Job: Industries |  |  |  |
| Engineering and construction<sup>1</sup> | 460 (6.7) | 3,500 (5.4) | 3,960 (5.5) |
| Textile and Cosmetics<sup>2</sup> | 538 (7.8) | 3,441 (5.3) | 3,979 (5.6) |
| Manufacturing | 897 (13.0) | 6,533 (10.1) | 7,430 (10.4) |
| Trading and Mass media<sup>3</sup> | 204 (3.0) | 1,895 (2.9) | 2,099 (2.9) |
| Distributors, Retailers | 318 (4.6) | 3,280 (5.1) | 3,598 (5.0) |
| Carriers<sup>4</sup> | 328 (4.7) | 2,802 (4.3) | 3,130 (4.4) |
| Public works | 407 (5.9) | 3,366 (5.2) | 3,773 (5.3) |
| IT industries<sup>5</sup> | 441 (6.4) | 2,942 (4.6) | 3,383 (4.7) |
| Banks and Financial services | 206 (3.0) | 1,868 (2.9) | 2,074 (2.9) |
| Food and Other Services<sup>6</sup> | 722 (10.4) | 7,190 (11.1) | 7,912 (11.1) |
| Medical care, Welfare | 469 (6.8) | 4,754 (7.4) | 5,223 (7.3) |
| Education | 235 (3.4) | 2,354 (3.6) | 2,589 (3.6) |
| Others<sup>7</sup> | 473 (6.8) | 4,323 (6.7) | 4,796 (6.7) |
| Not applicable<sup>8</sup> | 1,213 (17.6) | 16,276 (25.2) | 17,489 (24.5) |
| **Exposures** |  |  |  |
| Have a PS5 (yes) | 2,088 (30.2) | 928 (1.4) | 3,016 (4.2) |
| Played PS5 this month (yes) | 1,330 (19.2) | 395 (0.6) | 1,725 (2.4) |
| Video game play time |  |  |  |
| <1 hour/day | 3,632 (52.6) | 48,930 (75.8) | 52,562 (73.6) |
| 1-3 hours/day | 2,801 (40.5) | 13,386 (20.7) | 16,187 (22.7) |
| More than 3 hours/day | 478 (6.9) | 2,208 (3.4) | 2,686 (3.8) |
| Smartphone game play time |  |  |  |
| <1 hour/day | 3,679 (53.2) | 39,219 (60.8) | 42,898 (60.1) |
| 1-3 hours/day | 2,814 (40.7) | 22,224 (34.4) | 25,038 (35.1) |
| More than 3 hours/day | 418 (6.0) | 3,081 (4.8) | 3,499 (4.9) |
| Win PS5 lottery (yes) | 2,449 (35.4) |  | 2,449 (35.4) |

Notes. PS5, PlayStation5. Respondents' characteristics are displayed. Caregivers' characteristics are used where appropriate.
<sup>1</sup>Civil engineering, Construction, Real estate, Housing and building services; <sup>2</sup>Daily necessities, Textile and apparel, Cosmetics, Food and Beverages; <sup>3</sup>Trading companies, Publishing, Printing, Mass media; <sup>4</sup>Carriers, Warehousing, Logistics; <sup>5</sup>Software and Information services; <sup>6</sup>Food services, Hairdressing, Cosmetology, Other Services; <sup>7</sup>Other industries and types of business; <sup>8</sup>Not applicable (including no answer)

### Multivariate regression and PSM

The ITT effects of winning game console lotteries, estimated using multivariate regression and PSM approach, are illustrated in Fig 1. Winning game console lotteries improved mental health by 0.07 SD (P<0.05, regression) and 0.03 SD (P=0.17, PSM), and life satisfaction by 0.11 SD (P<0.01, regression) and 0.11 SD (P<0.01, PSM). All estimates were statistically significant except for the PSM-estimated effect on psychological distress. Note that the negative effect on K6 indicates reductions in psychological distress.

**Figure 1:**
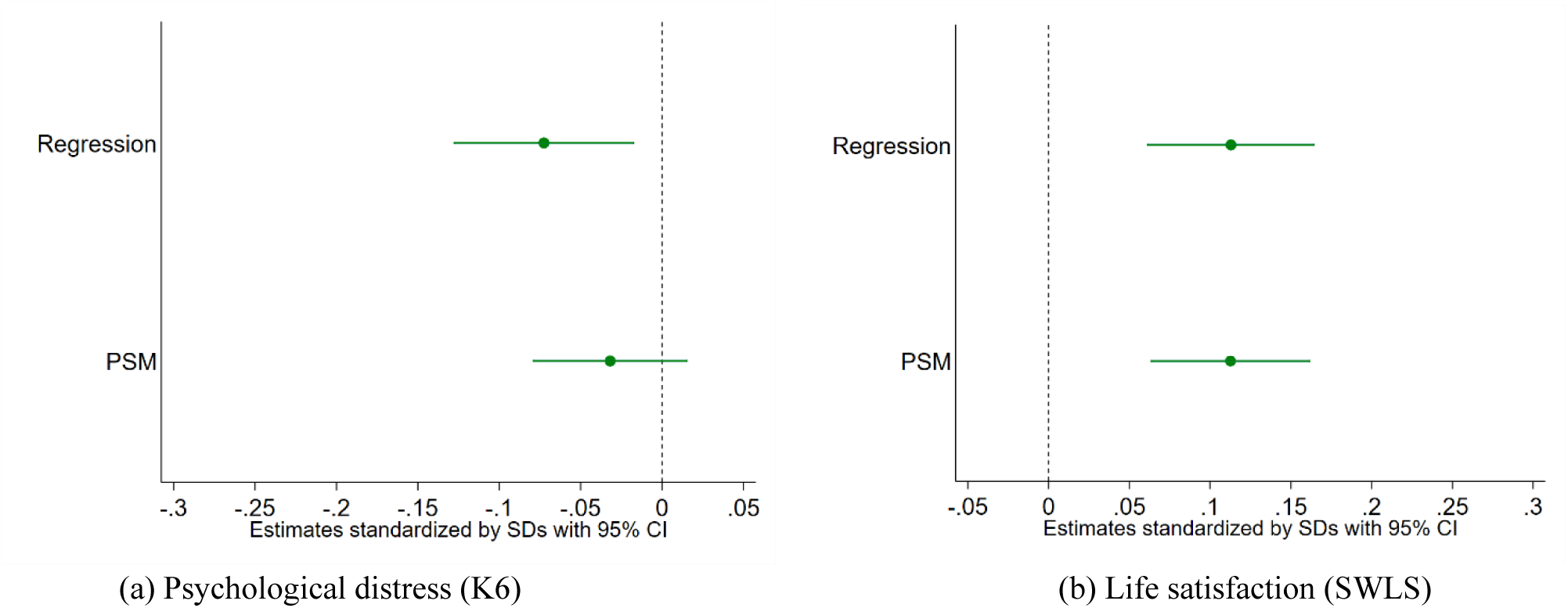
Causal effects on mental well-being from winning PS5 lotteries in post-COVID Japan (N=6,911). Notes: CI, confidence intervals; PSM, Propensity Score Matching; PS5, PlayStation5. The analysis sample is limited to those who joined PS5 lotteries. The point estimates and the 95 percent confidence intervals are shown. Regression standard errors are clustered by prefectures. Abadie-Imbens robust standard errors are used for PSM. Equation (1) is used for the regression estimates (eMethods). A lower K6 means having less psychological distress, while a higher SWLS means greater life satisfaction. The estimates are standardized by the standard deviations.

Additionally, lottery winners played video games 0.23h (P<0.01, regression; 0.23h, P<0.01, PSM) longer per day than non-winners and decreased smartphone gameplay duration by 0.09h (P<0.01, regression; 0.08h, p=0.018, PSM) (Supplementary Fig 2).

### Assessing natural experiment validity

The background characteristics of the lottery winners and non-winners were similar (Supplementary Table 5). As expected, only 1 of 30 variables exhibited standardized differences exceeding 0.20 in absolute value: the number of times that respondents joined the lotteries. In addition, the effects on pseudo-outcomes were small and not statistically significant, supporting the plausibility of conditional exchangeability (Supplementary Table 6; further discussion in eResults). Moreover, covariate balance after matching and common support showed that our PSM analysis achieved balance between the treatment and control groups (Supplementary Fig 3 and Supplementary Fig 4).

### IV approach

The local average treatment effects (LATE) of owning PS5 on mental well-being, estimated via the IV method, are shown in panel a of Fig 2. Possessing PS5 improved mental health by 0.13 SD and enhanced life satisfaction by 0.20 SD. Past-month PS5 play led to 0.21 SD improvements in mental health and 0.32 SD improvements in life satisfaction. An extra hour of daily video game play resulted in 0.32 SD improvements in mental health and 0.50 SD improvements in life satisfaction. Possession of PS5 increased video gaming duration by 0.40 h (Supplementary Fig 5). Weak instrument tests detected no issues (Supplementary Table 7 and Supplementary Table 8).

**Figure 2:**
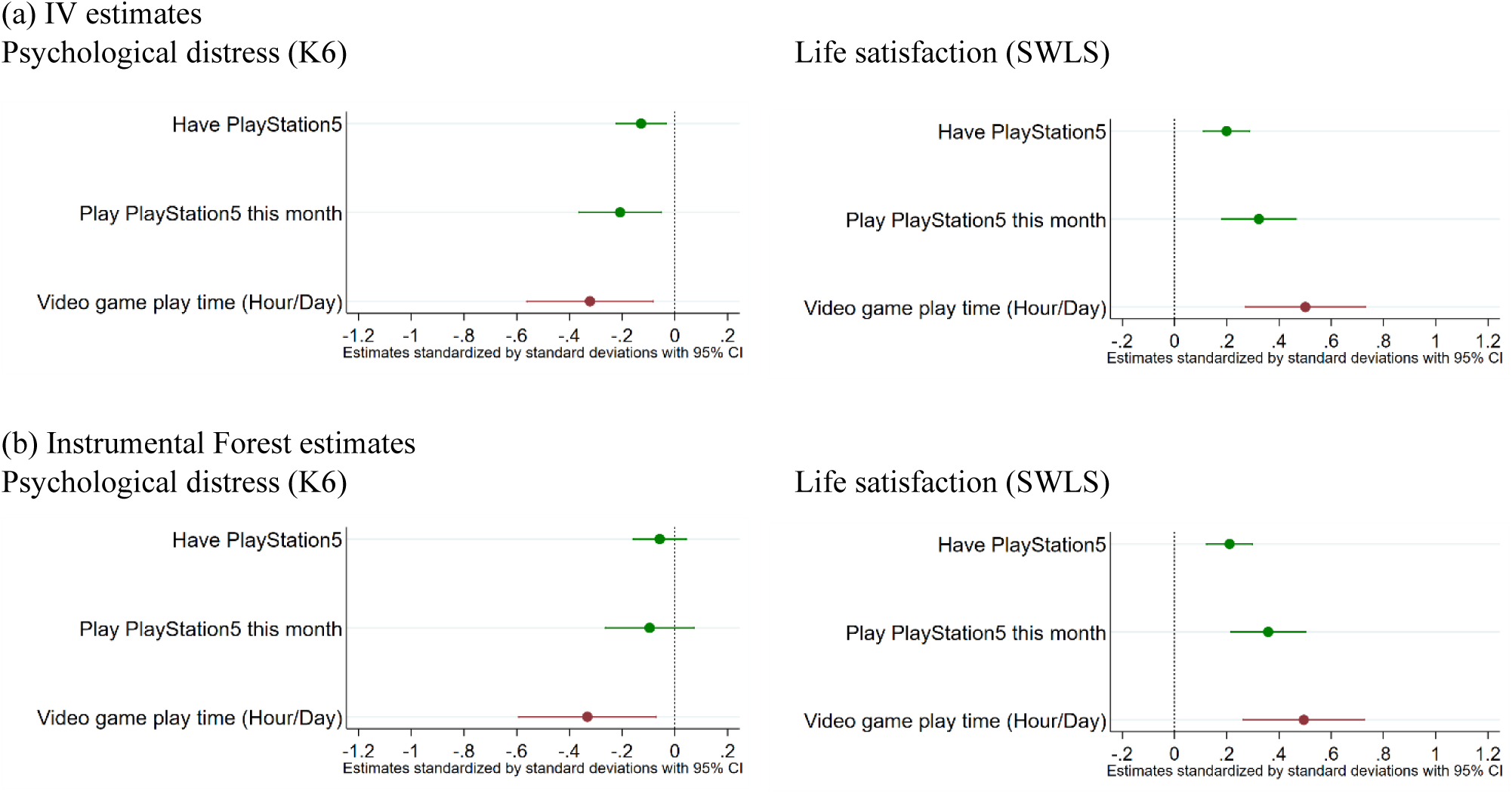
Causal effects of video game engagement on mental well-being in post-COVID Japan (N=6,911). Notes: CI, confidence intervals. The causal effect of video game engagement on mental well-being is estimated by the IV method and instrumental forests. The outcomes are psychological distress and life satisfaction. The analysis sample is limited to those who joined PS5 lotteries. The point estimates (mean values) and the 95% CIs are shown. Standard errors are clustered by prefectures. A lower K6 score indicates less psychological distress, whereas a higher SWLS score indicates greater life satisfaction. The estimates are standardized by the SD. The IV estimates are also presented in Supplementary Table 9. The estimates for possession of video game consoles are preferred as they are more likely to adhere to the exclusion restriction requirement, a nontestable assumption of the IV method. The estimates based on console usage and play duration are more prone to violating this assumption.

These estimates are close to those in Egami et al.^30^ For example, the during-COVID study’s estimates of the psychological effects of owning PS5 were 0.12 SD improvements in mental health and 0.23 SD improvements in life satisfaction. Fig 3 illustrates a comparison of the mental effects between the during-COVID study and the current post-COVID study.

**Figure 3:**
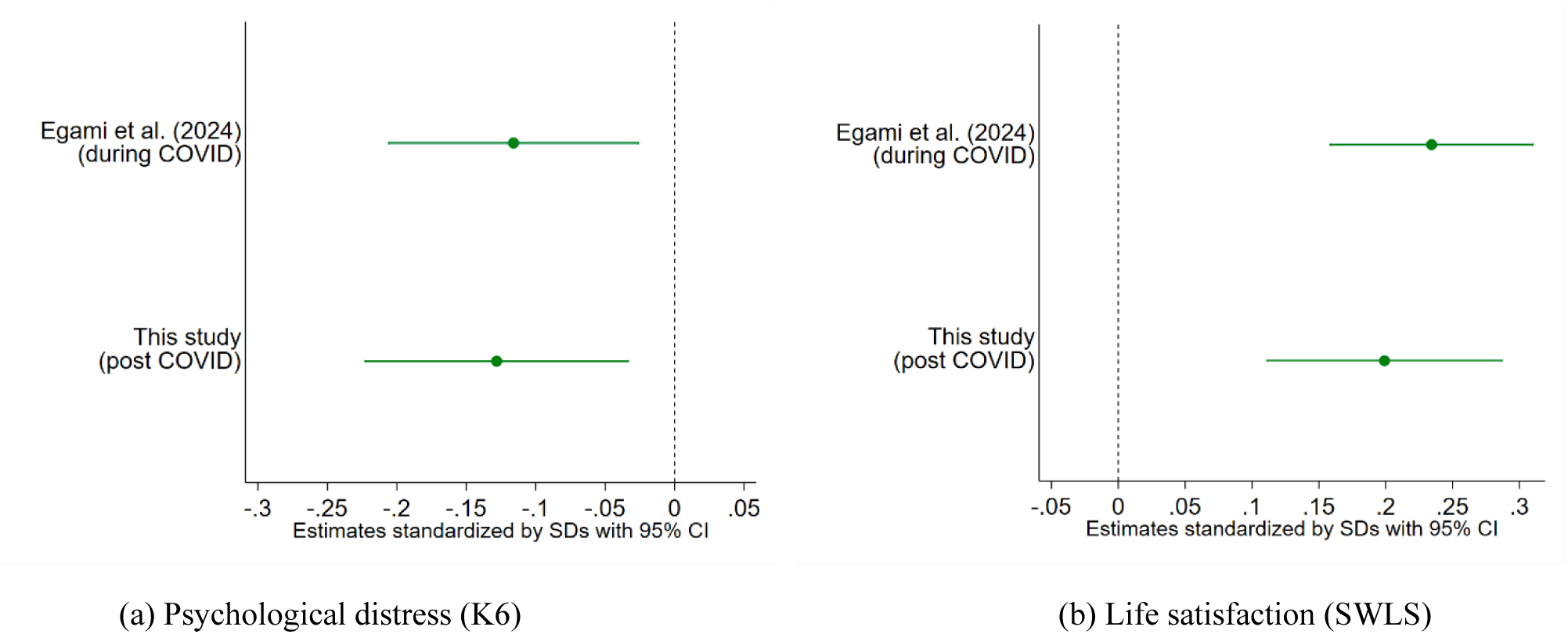
Comparison between effects of video game console ownership on mental well-being during the COVID-19 and post-COVID-19 period (N=6,911). Notes: CI, confidence intervals; IV, Instrumental Variable. The effects on psychological distress (a) and on life satisfaction (b). The effect of possession of PlayStation 5 on mental well-being is estimated by the IV method. The estimates from Egami et al. and this study are shown. The analysis sample is limited to those who joined game console lotteries. The point estimates (mean values) and the 95% CIs are shown. Standard errors are clustered by prefectures. A lower K6 score indicates less psychological distress, whereas a higher SWLS score indicates greater life satisfaction. The estimates are standardized by the SD. Other IV estimates are presented in Supplementary Table 9 and Supplementary Fig 10.

Subgroup IV analysis (exposure: past-month PS5 play) in Supplementary Fig 6 demonstrated that the subgroup with longer gaming duration had a smaller mental benefit. The largest benefits were observed in the subgroups that played video games for about an hour.

### Machine learning: Instrumental forest

The LATE (or ACLATE) of game engagement on mental well-being, estimated via the Instrumental Forest, is shown in panel b of Fig 2. PS5 ownership improved life satisfaction by 0.21 SD, and past-month PS5 play enhanced life satisfaction by 0.36 SD. The corresponding effects on psychological distress were 0.06 SD and 0.10 SD, respectively, though neither was statistically significant. An extra hour of daily video game play resulted in 0.33 SD improvements in mental health and 0.50 SD improvements in life satisfaction. A weak instrument issue was not found.

### Sensitivity Analysis

The IV estimates from the sensitivity analysis, using data excluding round 6, were close to those from our main analysis and supported a mental benefit (Supplementary Fig 7). Possessing PS5 improved mental well-being (0.14 SD on the K6 and 0.18 SD on life satisfaction). Past-month PS5 play also led to enhanced mental well-being (0.22 SD on the K6 and 0.29 SD on life satisfaction). An extra hour of daily video game play improved mental well-being (0.34 SD on the K6 and 0.44 SD on life satisfaction).

## Discussion

This study examines the causal relationship between video gaming and mental well-being within a real-world context, particularly in the post-COVID period. Our natural-experimental approach demonstrated that winning a lottery for PS5 had a positive effect on mental well-being among individuals aged 10-69 years in Japan, via multivariate regression and PSM methods. The IV method showed that engagement with video games—PS5 ownership, past-month PS5 play, and video gaming duration—positively impacted mental well-being. These findings were robust to sensitivity analyses and were corroborated by machine learning estimates. Results from this multifaceted analytical approach consistently pointed to a positive effect of video gaming on mental well-being.

Our study illustrated a positive causal effect (ITT) of winning game console lotteries on mental well-being. Both multivariate regression and PSM indicate reductions in psychological distress and improvements in life satisfaction. Although one estimate is not statistically significant (the PSM-estimated effect on psychological distress), the direction remains consistent. It should be noted that our approach carries a lower risk of confounding bias than association analyses using game engagement indicators (e.g., gaming duration) as the exposure. Lottery winners also played video games longer, consistent with increased engagement as the underlying mechanism. While smartphone gaming duration declined among winners, video gaming duration increased by twice as much.

IV estimates indicated a positive causal effect (LATE) of PS5 ownership and past-month PS5 play on mental well-being, ranging from 0.1 to 0.3 SD, with larger effect sizes when past-month PS5 play was used as the endogenous variable. LATE estimates from the instrumental forest algorithm were consistent with these findings, although some estimates of psychological distress were not statistically significant.

An extra hour of daily video game play resulted in 0.3-0.5 SD improvements in mental well-being, though these effects were attenuated in subgroups with longer gaming hours. Because the IV estimates are based on video gaming among individuals who bought PS5 after winning the lottery, with a 0.40-hour increase in gaming duration, the findings might not generalize directly to substantially different settings (further discussed in eResults).

A multi-faceted assessment supports the credibility of our natural experiment: background characteristics were similar between lottery winners and non-winners, except for the (expected) number of lottery entries — reasonable, since more entries mean a greater propensity to win — which we addressed by including it as a covariate. Pseudo-outcome analysis supported unconfoundedness, and PSM validity checks (covariate balance and common support) confirmed adequate matching quality.

The estimates of video gaming’s effect on mental well-being obtained in this study are close to those reported in Egami et al.^30^ Egami et al. analyzed data collected during the COVID-19 pandemic, between March 2021 and March 2022, whereas this study used data collected between September 2022 and March 2023. The similarity of estimates across these two studies suggests that gaming’s effect on mental well-being remained stable across the COVID and post-COVID periods. However, this similarity is inconsistent with the expectation that gaming’s benefits were larger than usual during the pandemic,^30^ given that limited outdoor exercise has often been cited as a major contributor to psychological distress during this period. One possible explanation is that our analytical sample might mainly consist of individuals who were less inclined to outdoor activities, regardless of stay-at-home requirements. If so, their behavior may have changed little between the pandemic and post-COVID periods. Moreover, it remains empirically untested whether people would engage in outdoor activities in the absence of video gaming.^41^ Outdoor exercise might be a less common alternative to gaming than is often assumed.^23^

The positive effect of gaming on mental well-being could result from positive mechanisms that outweigh negative ones. Positive mechanisms include facilitating social interactions by initiating connections between users, while negative mechanisms include displacement of more psychosocially beneficial activities (e.g., sleep).^31^ Both mechanisms have been thoroughly explored in the literature, but few have been empirically tested using a method interpreted as causal.

Our sensitivity analysis, excluding the September 2022 data, yielded estimates close to our main results. This suggests that our data period, which is relatively close to the COVID-19 pandemic, is not a critical concern.

Although this study enriches the empirical evidence on diverse contexts of digital media use, external validity remains an important consideration. Estimates may differ in periods further removed from the pandemic and are likely to vary across gaming platforms, genres, populations, and social contexts. As with most lifestyle-effect research, continued accumulation of evidence across diverse settings is needed for a full picture of digital media effects.

### Strengths and limitations

This study has several strengths: a natural experiment approach with multifaceted robustness checks, and stronger external validity than laboratory studies given its use of real-world data from large samples (individuals aged 10–69 across all 47 prefectures). It also has limitations. First, the response rate was relatively low (46.6%), though this is typical of online surveys (averaging 44%^42^); we used PSM to mitigate potential bias from this. Second, as in most lottery studies, data were not collected separately for each lottery round, which would have enabled more direct comparisons between groups.^30^ Finally, gaming behaviour and well-being were self-reported. However, randomization in a natural experiment alleviates concerns about self-reporting bias, since lottery assignment is unlikely to correlate with reporting behaviour.

## Conclusion

This natural experiment found that video gaming improved mental well-being in the post-COVID period. The unexpectedly similar effect size relative to a related COVID-period study^30^ strengthens the cross-temporal robustness of our findings and enriches the evidence base on digital media and mental health. Our findings add to the growing evidence that digital media screen time has diverse effects on well-being and highlight the mental benefit of moderate gaming.

## Ethics Statement

This study complies with all relevant ethical regulations for research involving human participants. The survey was approved by the institutional review board of Takasaki City University of Economics (approval number 245-1). Informed consent was obtained from all participants by the survey firm before the interview. All data were kept confidential and used only for research purposes. The study posed minimal risk to participants, and their privacy was protected throughout the study. Data were anonymized to protect the participants’ privacy. Although the survey firm compensated the participants, the details of this payment were not disclosed to the research team. This study was not pre-registered.

## Acknowledgments

We thank Y. Yoshinari for technical and graphical support; H. Kanemitsu and T. Kinoshita for precious feedback on our survey questionnaire; and GRI for conducting omnibus surveys together with us and sharing invaluable data.

## Data availability

Data analyzed in this study were collected through a collaborative arrangement with GRI and Cross Marketing Inc. The data cannot be made publicly available due to contractual and legal restrictions on data use, participant consent, privacy considerations, and the commercially sensitive nature of these survey data. To support verification of the analytic code and transparency, synthetic data generated using the SDV library have been deposited on GitHub (https://github.com/kanigeruge/gamestudy-postcovid). However, the data are available upon request by accredited academic researchers from the corresponding author and with permission from GRI and Cross Marketing Inc., subject to applicable contractual and ethical restrictions.

## Code availability

In this study, we utilized standard methods and did not rely on custom code or specialized mathematical algorithms. The code associated with the major analyses presented in this manuscript (software: Stata 16.1, R version 4.5.2 with R package ‘grf’, version 2.6.0) is found on GitHub (https://github.com/kanigeruge/gamestudy-postcovid).

## Author contributions

H.E., M.S.R., T.Y., C.E. and T.W. had full access to all of the data in the study and took responsibility for the integrity of the data and the accuracy of the data analysis. Concept and design: H.E., T.Y., C.E. and T.W. Drafting of the manuscript: H.E., M.S.R., T.Y. and C.E. Critical revision of the manuscript for important intellectual content: M.S.R., S.H., and A.K.P. Statistical analysis: H.E., T.Y., C.E., and S.H. Obtained funding: H.E., T.Y., C.E. and T.W. Administrative, technical or material support: H.E., T.Y., C.E. and T.W. Supervision: H.E., M.S.R., and A.K.P.

