## Supplementary Information for "Causal effect of video gaming on mental well-being in Post-COVID Japan"

**Contents**


- **Supplementary Methods.**
    - eMethod 1. Additional information on lottery, sampling process, and data collection.
        - eMethod 1.1. Lotteries for PlayStation5.
        - eMethod 1.2. Sampling process.
        - eMethod 1.3. Lottery questionnaire.
        - eMethod 1.4. Omnibus online surveys.
    - eMethod 2. Additional details on statistical analysis.
        - eMethod 2.1. Causal assumptions.
        - eMethod 2.2. Reduced form analysis (multivariate regression) and PSM.
        - eMethod 2.3. Assessing natural experiment validity.
            - Placebo analysis (pseudo-outcome tests).
            - Methodological challenge in lottery-based natural experiment.
        - eMethod 2.4. Instrumental variable (IV) approach.
        - eMethod 2.5. Machine learning details.
- **Supplementary Results.**
    - eResult 1. Balance table.
    - eResult 2. Assessing natural experiment validity.
        - Assessment of balance and common support in PSM analysis.
        - Placebo analysis.
    - eResult 3. Comparison between effects of video gaming on mental well-being during the COVID-19 and post-COVID-19 period.
    - eResult 4. IV estimates and generalizability of findings.
- **Supplementary Tables.**
    - Supplementary Table 1: Number of observations for each round.
    - Supplementary Table 2: Exposures: variables capturing game engagement.

  - Supplementary Table 3: Description of covariates.
  - Supplementary Table 4: Video gaming preference and video game play duration among respondents.
  - Supplementary Table 5: Baseline characteristics' balance table for lottery winners and non-winners.
  - Supplementary Table 6: Assessing unconfoundedness for PS5 lottery data: pseudo outcomes.
  - Supplementary Table 7: Weak instrument tests.
  - Supplementary Table 8: First-stage regression results of instrumental variable method.
  - Supplementary Table 9: Causal impact of video game engagement on well-being in Japan.
  - Supplementary Table 10: List of covariates used as features in causal forest algorithm.
  - Supplementary Table 11: Outcomes by exposures.
  - Supplementary Table 12: Sensitivity check: Causal impact of video game engagement on well-being in Japan.
- **Supplementary Figures.**
  - Supplementary Figure 1: Survey participants and analysis sample.
  - Supplementary Figure 2: Causal effect of winning game console lotteries on gaming duration (N=6,911).
  - Supplementary Figure 3: Balance check before and after propensity score matching for estimations.
  - Supplementary Figure 4: Common support check for propensity score matching estimations.
  - Supplementary Figure 5: Causal effect of ownership of PS5 on gaming duration estimated by instrumental variable method (N=6,911).
  - Supplementary Figure 6: Subgroup analysis of treatment effect using instrumental variable method.
  - Supplementary Figure 7: Sensitivity check: Causal effects of video game engagement on mental well-being in post-COVID Japan (N=4,271).
  - Supplementary Figure 8: Price history of PS5 (Japanese Yen).
  - Supplementary Figure 9: Survey schedule and COVID-19 situation.
  - Supplementary Figure 10: Comparison between effects of video game engagement on mental well-being during the COVID-19 and post-COVID-19 period (N=6,911).
- **References.**

**Supplementary Methods.**

**eMethod 1. Additional information on lottery, sampling process, and data collection.**

*eMethod 1.1. Lotteries for PlayStation5.*

Since the initial release in 2020, PlayStation 5 (PS5) consoles have achieved widespread global recognition. By 2022, global sales of the PS5 had reached 10 million. The COVID-19 pandemic further boosted player numbers, with consoles serving as communication platforms for connecting with friends and family remotely.[1] However, there was a semiconductor shortage that hindered the general public in Japan from purchasing the PS5 through standard channels, such as online or in-store shopping. Instead, most consumers had to enter lotteries run by retailers to acquire the consoles. One could alternatively explore the secondary market to buy a second-hand console by paying an additional cost. See Supplementary Fig 8 for the price history of the PS5.

It is worth describing the context of the PS5 lotteries because it relates to the unconfoundedness of the lottery results (further details are provided in Egami et al.[2]). Customers were first required to register for a PS5 lottery. When registering, participants were required to provide personal information, including their name and email address, via the retailer's website or in person. Although some retailers required membership in their clubs to participate in the lottery, membership was accessible to everyone. Retailers did not target certain customers with the private information collected, such as gender or age. Once an individual won a lottery, they gained the privilege of purchasing a PS5.

Participants might enter multiple lotteries concurrently, often with assistance from family members. For example, an individual intensely desiring a PS5 was able to participate in lotteries offered by various retailers and involve family members. One of our control variables representing the number of lottery participations included those of family members. In addition, the announcement of lottery results did not follow a consistent timeline; retailers lacked a uniform schedule for lotteries. The waiting period fluctuated between two weeks and several months. Non-winning participants could not immediately enter the next lottery and had to wait until new ones were announced; meanwhile, participants were free to enter another lottery offered by a different retailer.

*eMethod 1.2. Sampling process.*

This study's survey sample size (see Supplementary Table 1) varies by round depending on the business objective of the research firm—gameage R&I (GRI); the research firm conducted surveys with support from the survey firm Cross Marketing. GRI aimed to collect a specific number of opinions from individuals who played particular gaming software, to sell this information to its customers, who are usually video game companies. Therefore, if the number of people who played a gaming software was small, GRI had to collect more data. Conversely, if more respondents played the gaming software, GRI was able to stop collecting data after collecting a smaller sample. Namely, the sample size was determined by GRI's business objective, and the authors did not use statistical methods to predetermine sample sizes.

GRI used stratified random sampling (stratified by gender, age, and gaming preference). Yet the actual number of responses in each stratum is not equal. This is because GRI's business included selling information about video game players who played certain gaming software; they had to collect more responses from individuals who played video games. As a result, our sample tends to include fewer older adults.

Participants were blind during the survey. The survey firm (Cross Marketing) conducted some data cleaning before the authors received the data; they removed specific observations, including those with too many missing answers, among other low-quality indicators—a process to which the authors were blind. Upon initiating data analysis, the authors were unblinded. Although Cross Marketing compensated the respondents, the details of their payment were not disclosed to the authors.

*eMethod 1.3. Lottery questionnaire.*

We used two types of questions to collect information on PS5 lotteries. First, we asked respondents how many times they (including their household members and relatives) entered lotteries to buy PS5s for their households. We summed these entries and used the result to create a variable indicating the number of times respondents joined the lotteries. Second, we asked about the retailers (for example, Sony Store, Yodobashi Camera, and Amazon) where they participated in the lotteries. We calculated the total number of times each household joined the lotteries by summing the counts for each retailer. As expected, the two questions yielded similar total numbers of lottery participations. We aimed to help respondents accurately recall their lottery participation. Our analysis used the first item. Some respondents reported that they had not yet been informed of the lottery results, and we coded them as having "not win lottery=lose lottery."

#### *eMethod 1.4. Omnibus online surveys.*

GRI, a gaming market research firm, sent surveys to individuals via collaboration with a survey firm, Cross Marketing. The two companies collaborated to construct and maintain a panel of registered survey respondents. Their purpose was to collect behavioral data from video game users as well as non-users. Our additional questions were incorporated occasionally. This study used data collected in September, December 2022, and March 2023 (hereafter Rounds 6-8). Egami et al. used data collected in December 2020, March, May, November 2021, and March 2022 (hereafter Rounds 1-5).

Our data are a blend of panel and repeated cross-sectional observations. For instance, 38.6% (34,627/89,742) of the respondents answered our survey more than once during Rounds 2 to 8. This data structure facilitates placebo analysis.

### eMethod 2. Additional details on statistical analysis.

#### *eMethod 2.1. Causal assumptions.*

The causal inference method in this study is based on the potential outcomes framework. We rely on three key assumptions — standard in empirical research — that must hold: conditional unconfoundedness (also known as conditional exchangeability, conditional mean independence, or unconfoundedness), positivity, and consistency.[3] The causal assumptions required for this study are the same as those in Egami et al.[2] While we explain the essence below, further details are available in Egami et al.[2]

*Conditional exchangeability*: Conditional exchangeability is expressed as follows:

$$(Y_{0i}, Y_{1i}) \perp D_i \mid X_i$$

where $Y_{0i}$ and $Y_{1i}$ represent the potential outcomes for a treatment variable $D_i$, and $X_i$ is a set of observed covariates.

Conditional exchangeability is ultimately untestable, yet a natural experimental design enhances its plausibility. To evaluate this, this study examined the covariate balance between the treatment and control groups. As recommended in the literature,[4] we also used pseudo-outcome tests to assess the plausibility of conditional exchangeability.

*Positivity*: Positivity is assessed using the common support check figures prepared for the propensity score matching (PSM). There is a good overlap between the treatment and control groups thanks to the natural experiment.

*Consistency*: As in a standard empirical observational study, it is reasonable to assume consistency because the treatment is well-defined: winning a PS5 lottery. As for the instrumental variable (IV) method, exposure variables such as possession of PS5, played PS5 last month, and video gaming time are also well-defined. Yet, reservations should be made for potential violations. For example, different versions of treatment may exist as PS5 has multiple video game software.[3]

In the multivariate regression approach, we adjusted for the number of times the respondent household participated in the lotteries. We assume that the results of the game console lotteries are random, given the number of times respondents participated in the lotteries. We also considered geographical differences between respondents by controlling for prefecture (dummy variables), as where they joined a PS5 lottery might have mattered. Those basics are common to all the methods we used in this study.

*eMethod 2.2. Reduced form analysis (multivariate regression) and PSM.*

We used the following equation to conduct an Intention-to-Treat (ITT) analysis:

$$Y_{ip} = \beta Lottery_{ip} + \psi X_{ip} + \phi Join_{ip} + \alpha_p + \epsilon_{ip} \quad (1)$$

where $Y_{ip}$ is an outcome variable such as a well-being measure for an individual $i$ in prefecture $p$. $Lottery_{ip}$ is a dummy variable taking the value 1 if an individual wins the lottery and 0 otherwise. The coefficient $\beta$ is the parameter of interest. $X_{ip}$ is a set of control variables consisting of individual characteristics, including age, gender, marital status, job status, and whether having children or not. We additionally include gaming preference as control variables: dummy variables for each category (hardcore gamer, core gamer, middle-core gamer, casual gamer, and non-gamer). $Join_{ip}$ is the number of times that the household of individual $i$ joined the lotteries. $\alpha_p$ are prefecture dummy variables.

PSM was additionally used to obtain ITT effect estimates. The empirical causal inference literature has acknowledged the limitations of depending exclusively on OLS linear regression in observational studies.[4–6] It is recommended to compare OLS estimates with those from other methods, including matching.[4,6] This approach assures that implications are not sensitive to the choice of estimators.

We implemented the PSM method following Imbens (2015)[4] and our previous work.[2] This study followed the design choices of our previous work, and more details can be found there.[2]

Logit models were used. The Stata command *psmatch2* was used to perform the matching. We also used the Stata command *psestimate* (whose code is based on Imbens's work[4]) to select the variables for matching without arbitrary variable choice. The selected variables, including the second-order terms, are found in Supplementary Fig 3. Consequently, observations with propensity scores less than 0.1 and more than 0.9 were trimmed. The propensity score for trimming was estimated using a design that involved one-to-one nearest-neighbor matching without replacement, in descending order.

After trimming the data, we conducted PSM analysis. To improve precision, a design using two-to-one nearest-neighbor matching with replacement was employed.[4,7]

*eMethod 2.3. Assessing natural experiment validity*

***eMethod 2.3.1 Placebo analysis (pseudo-outcome tests).***

We conducted a placebo analysis using previous surveys. Data collected between February 2021 and March 2022—survey rounds 2-5 as shown in Supplementary Table 1 and Supplementary Fig 9—were utilized.

We performed a placebo analysis (also known as a pseudo-outcome test) following Imbens (2015).[4] We used pre-lottery values for K6 and SWLS from rounds 2-5 as pseudo-outcomes, with earlier rounds prioritized. Thus, a round 6-8 dataset with pre-lottery outcomes (K6 and SWLS) was constructed. We applied ITT analyses, including regression and PSM, to the dataset.

To keep the sample clean, only individuals who could not win PS5 lotteries (lost or waited for results) in the corresponding past rounds were used when constructing pre-lottery values. Otherwise, the outcomes of lottery winners from past rounds were included as pseudo-outcomes.

***eMethod 2.3.2 Methodological challenge in lottery-based natural experiment.***

Natural experimental studies have advantages regarding internal validity. However, they are not completely the same as randomized experiments and thereby need to address inherent challenges. Firstly, even if a lottery randomly determines winners, a relatively low response rate, compared to a randomized experiment, poses a challenge. Systematic differences may exist within the subset of individuals who responded to the survey and are included as the analysis sample. Further, self-reported ticket purchasing behaviour could be another potential source of bias, a problem that does not arise in randomized experiments. Most importantly, most lottery studies are not even 'the ideal lottery study'[8] examining those who purchased the same number of lottery tickets in a given game. In the following paragraphs, we describe lottery-based natural experiments' methodological challenges. The current study's methodological challenges and underlying causal assumptions parallel those of our previous study and are discussed in detail there.[2]

While a randomized experiment gives every subject the same probability of receiving a treatment, lottery studies do not meet this criterion. A common approach to address this problem is to control for the factor of different probabilities of winning a lottery. Typically, the number of lottery tickets purchased is controlled as a covariate.

While controlling for the number of lottery tickets is useful, it does not completely solve methodological challenges stemming from differential lottery participation behavior, and thereby a lottery-based natural experimental study needs some assumptions. For example, previous studies (Imbens et al. and Doherty et al.)[8,9] treat different types of lotteries (or tickets) as identical in their regression analyses. In our study, we also had to treat various lotteries as equivalent, such as PS5 lotteries offered by different retailers.

When a study is not 'the ideal lottery study'[8] examining those who purchased the same number of lottery tickets in a given game, assuming a simple linear relationship for the number of lottery entries in a regression model may become problematic. Although we a priori know that the number of lottery entries should affect the propensity score, the precise functional form of this variable remains uncertain. In such scenarios, matching methods help address these methodological challenges.[4,6] Imbens (2015)[4] recommends using matching methods to strengthen findings from OLS regression analyses. He argues that matching methods can reduce reliance on the assumptions underlying OLS regression and mitigate potential bias arising from differential ticket-purchasing behaviour.

Our strategy to tackle the methodological challenges above is to incorporate PSM alongside regression analysis as recommended by Imbens (2015).[4] This reduces reliance on the assumption of a linear relationship for the number of lottery entries.[4,6] Also, we can better ensure that the matched groups are comparable in their observable characteristics (while a matching method does not address bias due to unobserved confounders). In this way, our approach also helps to lessen potential bias caused by non-response and self-reporting.

*eMethod 2.4. Instrumental variable (IV) approach.*

We examined various treatment variables, also known as exposure variables (or endogenous variables), to estimate the effect of gaming on mental well-being. We acknowledge that owning a PlayStation5 yields the most reliable causal inference, given that the exclusion restriction is plausibly unbreached. Owning a game console is the most fundamental step in experiencing the benefits or drawbacks of winning a game console lottery. On the other hand, the remaining variables are not free of exclusion-restriction issues. For instance, winning a PS5 lottery may not affect mental well-being solely through video gaming; prolonged web browsing from purchasing a PS5 may also have an impact. Thus, the exclusion restriction might be violated when using past-month PS5 play or video gaming duration as a treatment variable (particularly video gaming duration, which is not exclusive to PS5). Despite the potential violation, we present multiple IV models because they provide useful interpretation. For example, using video game duration as a treatment variable enables inference about how increasing play time affects mental well-being. We used a set of covariates, which are common to those used in the ITT analysis, to reduce the bias from potential confounders.

In addition, we conducted a subgroup IV analysis, using playing PS5 last month as the exposure variable. The subgroups were defined by gaming duration to explore differences in the game's psychological effects between those with short and those with long gaming durations. A caveat is that using a post-treatment variable, such as gaming duration, to define subgroups may introduce selection bias. Ideally, pre-treatment gaming duration data should be used, but such data were not available in our dataset.

*eMethod 2.5. Machine learning details.*

Generalized random forests (GRFs) are increasingly used to estimate treatment effects conditional on background characteristics for each individual (Conditional Average Treatment Effect,

CATE).[10–13] Previous studies,[14–16] including methodological guides,[17,18] support the reliability and usefulness of GRFs. However, the algorithm can also be used to estimate Average Treatment Effect (ATE). They use a doubly robust average treatment effect estimator (e.g., augmented inverse propensity weighting, AIPW) and provide a more accurate estimate than just averaging CATEs.

Among machine learning algorithms for estimating treatment effects, GRFs incorporate IV methods[19,20] essential for causal inference in the presence of imperfect compliance. The GRF includes a preprocessing step that encodes the problem structure, enabling the integration of IVs into the forest framework. To identify treatment effects, assumptions for the IV method must hold. Treatment effects conditional on individual characteristics estimated via IV are called Conditional Local Average Treatment Effects (CLATEs). The Local Average Treatment Effect (LATE) estimated via the GRF is referred to as the Average Conditional Local Average Treatment Effect (ACLATE).

Treatment effect estimation via IV using the GRF is referred to in multiple ways: IV forests,[19] instrumental forests,[21] or IV causal forests.[20] In this study, we refer to them as instrumental forests.

The instrumental forest was implemented using the R package "grf", version 2.6.0. Details of the process and selected hyperparameters are as follows. Before running instrumental forest, we estimated the effect of the features $X_i$ on $Y_i$, $W_i$, and $Z_i$ separately via three regression forests (orthogonalization). For each regression forest, the number of trees was set to 2,000. These forests provide centered outcomes that can be used in the following steps. We also conducted the first-stage estimation (i.e., estimating compliance scores), in which we estimated the effects of $Z_i$ on $W_i$ conditional on individual characteristics using a causal forest. As our main estimate, we grew an instrumental forest and estimated ACLATE, utilizing the compliance scores from the first-stage estimation. The hyperparameters of the causal forest and the instrumental forest were as follows: (i) the minimum leaf size was set to 50, (ii) the number of trees was set to 2000, (iii) the number of covariates considered at each split was set to $K/3$, with $K$ being the total number of predictors. The list of covariates is found in Supplementary Table 10. Cluster-robust estimation, using 47 prefectures as clusters, was employed.

### Supplementary Results.

#### eResult 1. Balance table.

Supplementary Table 5 presents descriptive statistics for the covariates among lottery winners and non-winners. Following the literature, we report standardized differences (or normalized differences).[4] The table shows minor differences, with only one of 30 standardized differences exceeding 0.2—a threshold considered small.[22] Given that the standardized difference cutoff for assessing balance after PSM is 0.1,[23] the characteristics are well balanced even before PSM is applied. This supports the unconfoundedness of the treatment, which is vital for interpreting our analysis's results as causal.

#### eResult 2. Assessing natural experiment validity.

##### *eResult 2.1. Assessment of balance and common support in PSM analysis.*

A common step in PSM analysis is to assess covariate balance and common support in the propensity score distribution. We assessed the balance of covariates included in the matching procedure and found no significant differences between the treated and control groups (Supplementary Fig 3). After matching, all standardized differences in covariates were smaller than 0.1, a common cutoff.[23] We examined the common support and observed no evidence of overlap issues (Supplementary Fig 4). These results showed that our PSM analysis successfully achieved balance.

##### *eResult 2.2. Placebo analysis.*

The placebo analysis provided support for the unconfoundedness assumption (Supplementary Table 6). As for pseudo-K6, Model 1 showed a negligible effect size and Model 2 showed a statistically nonsignificant effect in the opposite direction (relative to the effect on real K6). As for pseudo-SWLS, Model 2 showed a negligible effect size, whereas Model 1 showed a relatively large, though nonsignificant, effect size in the same direction as the effect on real SWLS. Given the number of comparisons across the placebo models and baseline characteristics (balance table), some variation could be expected even under unconfoundedness. Taken together with the baseline characteristics comparison and the propensity score matching diagnostics reported above, we view the overall body of evidence as reasonably supportive of the credibility of our causal estimates. Note that the sample size for the placebo analysis is small due to the nature of the data, which only partially have a longitudinal data structure.

**eResult 3. Comparison between effects of video gaming on mental well-being during the COVID-19 and post-COVID-19 period.**

Comparison of all the IV estimates between the effects of video gaming on mental well-being during the COVID-19 and post-COVID-19 periods is presented in Supplementary Fig 10, in addition to the comparison of our major IV estimates (exposure: ownership of PS5) displayed in Fig 2. This study's estimates are consistent with those of Egami et al., which used data from the COVID-19 period.[2]

**eResult 4. IV estimates and generalizability of findings.**

The generalizability of our findings to other contexts merits discussion. It is worth noting that our estimates are based on a particular group of respondents defined by the natural experimental study design. The IV estimates are drawn from video gaming among individuals who purchased a PS5 after winning the lottery and increased their gaming by an additional 0.40 hours. The sample was diverse, comprising both core gamers and non-gamers, but the findings may not directly apply to markedly different contexts. For instance, some observational studies that included only active gamers reported smaller effect sizes associated with an extra hour of daily video gaming on well-being.[24,25] Exposures also differ: the cited studies measured engagement with specific games, whereas our study captures general gaming behavior. Outcome measures differ as well: they used SPANE (Scale of Positive and Negative Experience) for well-being, whereas we use SWLS (Satisfaction with Life Scale). Such differences in sample and context may all contribute to divergent effect estimates. Regarding the effect of an extra hour of daily video gaming, the exclusion restriction also draws attention from a methodological perspective, because gaming's effect might not operate solely through changes in gaming time.

Using IV methods on subgroup samples with past-month PS5 play as the exposure, we additionally showed that the mental benefit was smaller for the longer-gaming subgroup (Supplementary Fig 6). The positive mental effects were highest among subgroups with 0.5-1.3 hours of gaming. This implies that gaming's effect is not monotonic with respect to gaming duration, consistent with previous studies.[26,27] Again, consideration of the methodological features of natural experiments and IV methods is important for interpreting the findings. They are based on an observed 0.40-hour increase in gaming time resulting from PS5 lottery wins. Therefore, this might not be directly generalized to a substantially different context. For example, it is questionable to generalize this to estimate the effect of (increased) gaming for 3 hours.

**Supplementary Tables.**

**Supplementary Table 1: Number of observations for each round.**

| Round | # of people that were sent survey offers | # of people that answered survey | # of people who joined lotteries | # of people who won lotteries | Switch lotteries or PS5 lotteries |
|---|---|---|---|---|---|
| 1 | 34,615 | 18,912 | 1,773 | 926 | Switch |
| 2 | 27,186 | 18,189 | 1,481 | 254 | PS5 |
| 3 | 29,462 | 18,479 | 1,447 | 332 | PS5 |
| 4 | 47,183 | 26,996 | 2,127 | 499 | PS5 |
| 5 | 26,143 | 15,026 | 1,364 | 312 | PS5 |
| 6 | 69,636 | 29,604 | 2,640 | 863 | PS5 |
| 7 | 47,401 | 24,151 | 2,515 | 934 | PS5 |
| 8 | 36,226 | 17,680 | 1,756 | 652 | PS5 |

Notes. PS5, PlayStation5. Switch, Nintendo Switch. The response rate of this study (R6~R8) was 46.6 percent (71,435/153,263). In total, 6,911 people joined the game console lotteries. The survey rounds shaded in gray are not used in this study; those were used in Egami et al[2].

**Supplementary Table 2: Exposures: variables capturing game engagement.**

| Variables | Source and notes |
|---|---|
| PS5 Ownership | Respondents were asked whether their household owned a PS5. A dichotomous variable is used in statistical analysis. |
| PS5 Usage | Respondents were asked whether they had played PS5 over the last 30 days. A dichotomous variable is used in statistical analysis. |
| Time spent playing video games | Time of video game play, including any video games on TV/computer, in hours per day. Respondents were asked how much time they spent playing video games on the weekdays (and, in another question, the weekends) over the past 30 days. |
| Time spent playing smartphone games | Time of playing smartphone games, in hours per day. Respondents were asked how much time they spent playing smartphone games on the weekdays (and, in another question, the weekends) over the past 30 days. |

Notes. PS5, PlayStation5. Supplementary Table 11 displays summary statistics of the outcome variables by exposures.

**Supplementary Table 3: Description of covariates.**

| Variables | Source and notes |
|---|---|
| Age | Age of respondents in years. |
| Gender | Male or female. In statistical analysis, the male gender was encoded as 1, while the female gender was encoded as 0. |
| Marital status | Married, unmarried, or divorced/separated. In statistical analysis, dichotomous variables were used, and each category was encoded as 1 and otherwise encoded as 0. |
| Having children | Yes or No. In statistical analysis, having children was encoded as 1 and otherwise 0. |
| Employment status | Full-time employee, part-time employee, student, stay-at-home wife/husband, unemployed, or self-employed/others. In statistical analysis, dichotomous variables were used, and each category was encoded as 1 and otherwise encoded as 0. |
| Video gaming preference | Hardcore gamers, Core gamers, Mid-core gamers, Casual gamers, or Non-gamers. The level of affection for the game declines in the order listed. In statistical analysis, each category was encoded as 1 and otherwise encoded as 0. The preference was assessed with a clustering algorithm by the research company GRI based on frequencies of game play and game software purchase, and categorized into five groups. The preference was measured in November 2019, where available. If not available, the one assessed in November 2020 or November 2021 was employed. This is to avoid bad control issues. |
| Number of times joining game console lotteries | The number of times respondent households joined PS5 lotteries were. To obtain estimates more robust to outliers, we winsorized (a statistical technique meaning replacement of outliers by less extreme values) the number of times joining lotteries at the top 1 percent (see, for example, Yale and Forsythe[28]). |
| Occupation | Occupation of respondents/caregivers. We categorized into 14 categories: 'Civil engineering, Construction, Real estate, Housing and building services', 'Daily necessaries, Textile and apparel, Cosmetics, Food and Beverages', 'Manufacturing', 'Trading companies, Publishing, Printing, Mass media', 'Distributors, Retailers', 'Carriers, Warehousing, Logistics', 'Public works', 'Software and Information services', 'Banks and Financial services', 'Food services, Hairdressing, Cosmetology, Other Services', 'Medical care, Welfare', 'Education', 'Other industries and types of business', and 'Not applicable/No answer.' Initially, the survey included 37 categories, which the authors reclassified. In statistical analysis, dichotomous variables were used, and every 14 types were encoded as 1 and otherwise encoded as 0. |
| Prefecture | Prefecture of residence. In statistical analysis, dichotomous variables were used, and every 47 prefectures were encoded as 1 and otherwise encoded as 0. |

Notes. GRI, gameage R&I.

**Supplementary Table 4: Video gaming preference and video game play duration among respondents.**

| Variable | Mean (SD) | | | | |
|---|---|---|---|---|---|
| | Hardcore gamer (n=11196) | Core gamer (n=17254) | Middle-core gamer (n=18964) | Casual gamer (n=17579) | Non-gamer (n=6442) |
| Video game play time (hour/day) | 1.358 (1.794) | 0.853 (1.334) | 0.488 (0.984) | 0.240 (0.743) | 0.099 (0.605) |
| Smartphone game playtime (hour/day) | 1.174 (1.654) | 1.070 (1.462) | 0.956 (1.309) | 0.595 (1.042) | 0.194 (0.782) |

Notes. Video gaming preferences were measured in November 2021 for round 6, and November 2022 for rounds 7 and 8. Whereas this table associates the most recent gaming preferences with gaming time for each survey round, our analyses, including balance checks, utilized the oldest available data, going back to 2019, to avoid bad control issues.

**Supplementary Table 5: Baseline characteristics' balance table for lottery winners and non-winners.**

| Variable | Did not win PS5 lottery (N=4462) Mean/(SD) | Won PS5 lottery (N=2499) Mean/(SD) | Normalized difference |
|---|---|---|---|
| Number of times joined lottery | 6.891 (11.610) | 9.664 (14.429) | -0.212 |
| Age | 37.805 (16.243) | 37.306 (15.635) | 0.031 |
| Gender (Male) | 0.615 (0.487) | 0.622 (0.485) | -0.015 |
| Married | 0.615 (0.487) | 0.618 (0.486) | -0.007 |
| Divorced/separated | 0.059 (0.236) | 0.062 (0.241) | -0.011 |
| Have child(ren) | 0.545 (0.498) | 0.526 (0.499) | 0.040 |
| Student | 0.223 (0.417) | 0.214 (0.410) | 0.022 |
| Stay-at-home wife/husband | 0.076 (0.265) | 0.079 (0.270) | -0.011 |
| Full-time employee | 0.474 (0.499) | 0.489 (0.500) | -0.031 |
| Part-time employee | 0.090 (0.286) | 0.083 (0.276) | 0.024 |
| Self employed/others | 0.072 (0.258) | 0.067 (0.249) | 0.020 |
| Unemployed/not a student | 0.065 (0.247) | 0.067 (0.251) | -0.010 |
| *Gaming Preference* | | | |
| Hardcore gamer | 0.297 (0.457) | 0.298 (0.458) | -0.003 |
| Core gamer | 0.281 (0.449) | 0.282 (0.450) | -0.003 |
| Middle-core gamer | 0.195 (0.396) | 0.200 (0.400) | -0.012 |
| Casual gamer | 0.096 (0.295) | 0.108 (0.310) | -0.038 |
| Non-gamer | 0.131 (0.337) | 0.112 (0.315) | 0.058 |
| *Job: industries* | | | |
| Engineering and construction[1] | 0.297 (0.457) | 0.298 (0.458) | -0.003 |
| Textile and Cosmetics[2] | 0.281 (0.449) | 0.282 (0.450) | -0.003 |
| Manufacturing | 0.195 (0.396) | 0.200 (0.400) | -0.012 |
| Trading and Mass media[3] | 0.096 (0.295) | 0.108 (0.310) | -0.038 |
| Distributors, Retailers | 0.131 (0.337) | 0.112 (0.315) | 0.058 |
| Carriers[4] | 0.069 (0.254) | 0.061 (0.240) | 0.033 |
| Public works | 0.078 (0.269) | 0.077 (0.266) | 0.006 |
| IT industries[5] | 0.132 (0.339) | 0.126 (0.332) | 0.019 |
| Banks and Financial services | 0.028 (0.164) | 0.033 (0.178) | -0.028 |
| Food and Other Services[6] | 0.050 (0.217) | 0.040 (0.195) | 0.048 |
| Medical care, Welfare | 0.048 (0.213) | 0.047 (0.212) | 0.001 |
| Education | 0.056 (0.229) | 0.065 (0.246) | -0.039 |
| Others[7] | 0.061 (0.239) | 0.069 (0.254) | -0.033 |

Notes. PS5, PlayStation5. Respondents' characteristics are displayed. Caregivers' characteristics are used where appropriate.

[1]Civil engineering, Construction, Real estate, Housing and building services; [2]Daily necessaries, Textile and apparel, Cosmetics, Food and Beverages; [3]Trading companies, Publishing, Printing, Mass media; [4]Carriers, Warehousing, Logistics; [5]Software and Information services; [6]Food services, Hairdressing, Cosmetology, Other Services; [7]Other industries and types of business.

**Supplementary Table 6: Assessing unconfoundedness for PS5 lottery data: pseudo outcomes.**

| Outcome variables | Pseudo Outcome (K6, Standardized) β coefficient (95% CI) | | Pseudo Outcome (SWLS, Standardized) β coefficient (95% CI) | |
|---|---|---|---|---|
| | Model 1: Regression | Model 2: PSM | Model 1: Regression | Model 2: PSM |
| Win PS5 lottery | 0.037 (-0.096 to 0.17) | -0.05 (-0.211 to 0.111) | 0.033 (-0.096 to 0.163) | 0.067 (-0.076 to 0.209) |
| Number of observations | 1,199 | 1,176 | 1,199 | 1,176 |

Notes. PS5, PlayStation5; K6, Kessler psychological distress scale; SWLS, The Satisfaction with Life Scale; PSM, Propensity Score Matching; CI, Confidence interval. Coefficients were standardized using standard deviation. Pseudo outcomes are K6 and SWLS measured at rounds 2-5, with earlier rounds prioritized. The sample consists of rounds 6-8 observations. Regressions were adjusted for a set of covariates (i.e., age, gender, marital status, employment status, whether having children or not, occupation, gaming preference, and the number of times joining PS5 lotteries) in Supplementary Table 3, prefecture dummies, and round dummies. Regression standard errors were clustered by prefectures. Abadie-Imbens robust standard errors were used for PSM.

**Supplementary Table 7: Weak instrument tests.**

| Variables | Number of observations | Kleibergen-Paap rk Wald F statistic | |
|---|---|---|---|
| | | Model 1 | Model 2 |
| ***Outcome variable: Psychological distress (K6)*** | | | |
| Have a PS5 | 6,911 | 2014 | 1867 |
| Played PS5 this month | 6,911 | 1325 | 1534 |
| Video game play duration (hour/day) | 6,911 | 57.44 | 33.09 |
| ***Outcome variable: Life satisfaction (SWLS)*** | | | |
| Have a PS5 | 6,911 | 2014 | 1867 |
| Played PS5 this month | 6,911 | 1325 | 1534 |
| Video game play duration (hour/day) | 6,911 | 57.44 | 33.09 |

Notes. PS5, PlayStation5. This table displays weak instrument test statistics corresponding to the models of the instrumental variable method in Supplementary Table 9. Each estimate used winning PlayStation5 lottery as its instrument. Exposure variables are presented in each row. Model 1: Adjusted for prefecture dummy variables and round dummy variables only. Model 2: Additionally adjusted for a set of covariates (i.e., age, gender, marital status, employment status, whether having children or not, occupation, gaming preference, and the number of times joining PS5 lotteries) in Supplementary Table 3. The threshold for detecting a weak instrument is <10.

**Supplementary Table 8: First-stage regression results of instrumental variable method.**

| Variables | Number of observations | Model 1 | | Model 2 | |
|---|---|---|---|---|---|
| | | $\beta$ coefficient (95% CI) | *P*-value | $\beta$ coefficient (95% CI) | *P*-value |
| ***Outcome variable: Have a PS5*** | | | | | |
| Win PS5 lottery | 6,911 | 0.568 (0.543 to 0.594) | <0.001 | 0.567 (0.54 to 0.593) | <0.001 |
| ***Outcome variable: Played PS5 this month*** | | | | | |
| Win PS5 lottery | 6,911 | 0.35 (0.331 to 0.37) | <0.001 | 0.349 (0.331 to 0.367) | <0.001 |
| ***Outcome variable: Video game play time (hour/day)*** | | | | | |
| Win PS5 lottery | 6,911 | 0.298 (0.219 to 0.378) | <0.001 | 0.225 (0.146 to 0.304) | <0.001 |

Notes. PS5, PlayStation5; CI, confidence intervals. This table displays first-stage regression results corresponding to the models in Supplementary Table 9. Model 1: Adjusted for prefecture dummy variables and round dummy variables only. Model 2: Additionally adjusted for a set of covariates, including gaming preferences. Standard errors were clustered by prefectures. The two-sided t-test was used as the statistical test. No adjustments for multiple comparisons were made.

**Supplementary Table 9: Causal impact of video game engagement on well-being in Japan.**

| Variables | Number of observations | Model 1 | | Model 2 | |
|---|---|---|---|---|---|
| | | $\beta$ coefficient (95% CI) | *P*-value | $\beta$ coefficient (95% CI) | *P*-value |
| ***Outcome variable: Psychological distress (K6)*** | | | | | |
| Have a PS5 | 6,911 | -0.078 (-0.186 to 0.029) | 0.152 | -0.128 (-0.223 to -0.033) | 0.009 |
| Played PS5 this month | 6,911 | -0.126 (-0.302 to 0.049) | 0.154 | -0.207 (-0.363 to -0.051) | 0.01 |
| Video game play duration (hour/day) | 6,911 | -0.148 (-0.348 to 0.051) | 0.143 | -0.322 (-0.56 to -0.083) | 0.009 |
| ***Outcome variable: Life satisfaction (SWLS)*** | | | | | |
| Have a PS5 | 6,911 | 0.199 (0.105 to 0.293) | <0.001 | 0.199 (0.11 to 0.287) | <0.001 |
| Played PS5 this month | 6,911 | 0.324 (0.172 to 0.475) | <0.001 | 0.323 (0.18 to 0.465) | <0.001 |
| Video game play duration (hour/day) | 6,911 | 0.38 (0.21 to 0.549) | <0.001 | 0.5 (0.271 to 0.73) | <0.001 |

Notes. PS5, PlayStation5; CI, confidence intervals; K6, Kessler psychological distress scale; SWLS, The Satisfaction with Life Scale. The instrumental variable method's estimates are presented. Each estimate used winning PlayStation5 lottery as its instrument. Exposure variables are presented in each row. Model 1: Adjusted for prefecture dummy variables and round dummy variables only. Model 2: Additionally adjusted for a set of covariates (i.e., age, gender, marital status, employment status, whether having children or not, occupation, gaming preference, and the number of times joining PS5 lotteries) in Supplementary Table 3. Standard errors were clustered by prefectures. The estimates were standardized by the standard deviations. The two-sided t-test was used as the statistical test. No adjustments for multiple comparisons were made.

**Supplementary Table 10: List of covariates used as features in causal forest algorithm.**

| Variable description | Type | Variable description | Type |
|---|---|---|---|
| Age | Continuous | Number of times joining game console lotteries | Continuous |
| Gender (Male) | Binary | **Number of lottery entries per household members** | |
| Have children (yes) | Binary | Respondent him/herself participated. | Continuous |
| **Marital status** | | Brothers/sisters participated. | Continuous |
| Married | Binary | Children participated. | Continuous |
| Divorced/separated | Binary | Parents/grandparents participated. | Continuous |
| Not married | Binary | Other siblings participated. | Continuous |
| **Occupation** | | **Number of lottery entries per stores** | |
| Student | Binary | Online lottery at Sony Store. | Continuous |
| Stay-at-home wife/husband | Binary | Online lottery at GEO. | Continuous |
| Full-time employee | Binary | Online lottery at Bic Camera. | Continuous |
| Part-time employee | Binary | Online lottery at Yamada Denki. | Continuous |
| Self employed/others | Binary | Online lottery at Joshin. | Continuous |
| Unemployed/not a student | Binary | Online lottery at Nojima. | Continuous |
| **Gaming preference** | | Online lottery at Aeon or Edion. | Continuous |
| Hardcore gamer | Binary | Online lottery at Don Quijote or Sofmap. | Continuous |
| Core gamer | Binary | Online lottery at other stores. | Continuous |
| Middle-core gamer | Binary | Over-the-counter lottery at any stores. | Continuous |
| Casual gamer | Binary | | |
| Non-gamer | Binary | | |
| **Job: Industries** | | | |
| Engineering and construction | Binary | | |
| Textile and Cosmetics | Binary | | |
| Manufacturing | Binary | | |
| Trading and Mass media | Binary | | |
| Distributors, Retailers | Binary | | |
| Carriers | Binary | | |
| Public works | Binary | | |
| IT industries | Binary | | |
| Banks and Financial services | Binary | | |
| Food and Other Services | Binary | | |
| Medical care, Welfare | Binary | | |
| Education | Binary | | |
| Others | Binary | | |
| Not applicable | Binary | | |
| Indicator of being answered by caregivers on behalf of children. | Binary | | |
| 47 Prefecture indicators for each | Binary | | |
| 3 round indicators for each | Binary | | |

Notes. As for the marital status (unmarried, married, divorced/separated) and the indicator of having children (yes, no), we employed two types of variables. One set of variables pertains to the attributes of either the respondents or caregivers, while the other set of variables relates to the respondents' characteristics.

**Supplementary Table 11: Outcomes by exposures.**

| | Psychological distress (K6) | | Life satisfaction (SWLS) | |
|---|---|---|---|---|
| | N | Mean (SD) | N | Mean (SD) |
| **Win PS5 lottery** | | | | |
| Yes | 2,449 | 6.9 (6.7) | 2,449 | 17.8 (6.8) |
| No | 4,462 | 7.1 (6.5) | 4,462 | 17.1 (6.5) |
| Total | 6,911 | 7.0 (6.6) | 6,911 | 17.4 (6.6) |
| **Have a PS5** | | | | |
| Yes | 3,016 | 5.8 (6.4) | 3,016 | 18.1 (6.8) |
| No | 68,419 | 5.7 (6.2) | 68,419 | 17.2 (6.4) |
| **Played PS5 this month** | | | | |
| Yes | 1,725 | 5.8 (6.4) | 1,725 | 18.1 (6.9) |
| No | 69,710 | 5.7 (6.2) | 69,710 | 17.2 (6.4) |
| Total | 71,435 | 5.7 (6.2) | 71,435 | 17.2 (6.4) |
| **Video game play time** | | | | |
| <1 hour/day | 52,562 | 5.7 (6.2) | 52,562 | 17.3 (6.4) |
| 1-3 hours/day | 16,187 | 5.8 (6.2) | 16,187 | 17.3 (6.5) |
| More than 3 hours/day | 2,686 | 6.7 (6.7) | 2,686 | 16.1 (7.0) |
| **Smartphone game play time** | | | | |
| <1 hour/day | 42,898 | 5.6 (6.2) | 42,898 | 17.4 (6.4) |
| 1-3 hours/day | 25,038 | 5.8 (6.1) | 25,038 | 17.2 (6.4) |
| More than 3 hours/day | 3,499 | 7.4 (6.7) | 3,499 | 16.0 (6.8) |
| Total | 71,435 | 5.7 (6.2) | 71,435 | 17.2 (6.4) |

Notes. PS5, PlayStation5.

**Supplementary Table 12: Sensitivity Check: Causal impact of video game engagement on well-being in Japan.**

| Variables | Number of observations | Model 1 | | Model 2 | |
|---|---|---|---|---|---|
| | | β coefficient (95% CI) | *P*-value | β coefficient (95% CI) | *P*-value |
| ***Outcome variable: Psychological distress (K6)*** | | | | | |
| Have a PS5 | 4,271 | -0.073 (-0.216 to 0.069) | 0.307 | -0.14 (-0.261 to -0.018) | 0.025 |
| Played PS5 this month | 4,271 | -0.118 (-0.348 to 0.112) | 0.309 | -0.224 (-0.42 to -0.028) | 0.026 |
| Video game play duration (hour/day) | 4,271 | -0.13 (-0.39 to 0.128) | 0.316 | -0.335 (-0.651 to -0.02) | 0.037 |
| ***Outcome variable: Life satisfaction (SWLS)*** | | | | | |
| Have a PS5 | 4,271 | 0.166 (0.054 to 0.277) | <0.001 | 0.183 (0.072 to 0.294) | <0.001 |
| Played PS5 this month | 4,271 | 0.266 (0.089 to 0.444) | <0.001 | 0.293 (0.117 to 0.47) | <0.001 |
| Video game play duration (hour/day) | 4,271 | 0.295 (0.09 to 0.5) | <0.001 | 0.44 (0.137 to 0.743) | <0.001 |

Notes. PS5, PlayStation5; CI, confidence intervals; K6, Kessler psychological distress scale; SWLS, The Satisfaction with Life Scale. The instrumental variable method's estimates are presented. Each estimate used winning PlayStation5 lottery as its instrument. Exposure variables are presented in each row. Model 1: Adjusted for prefecture dummy variables and round dummy variables only. Model 2: Additionally adjusted for a set of covariates (i.e., age, gender, marital status, employment status, whether having children or not, occupation, gaming preference, and the number of times joining game console lotteries) in Supplementary Table 3. Standard errors were clustered by prefectures. The estimates were standardized by the standard deviations. The two-sided t-test was used as the statistical test. No adjustments for multiple comparisons were made.

**Supplementary Figures.**

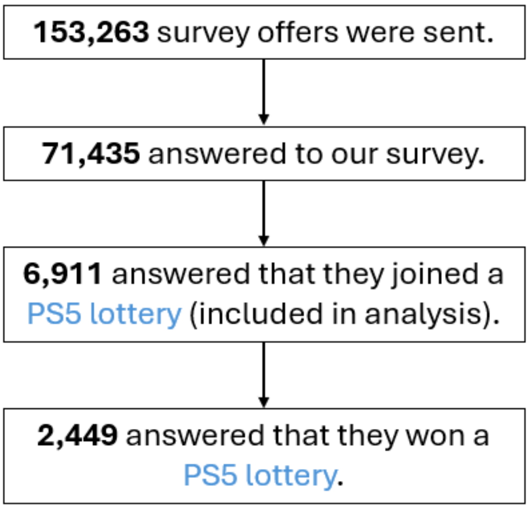


**Supplementary Figure 1: Survey participants and analysis sample.**
Notes. PS5, PlayStation5.

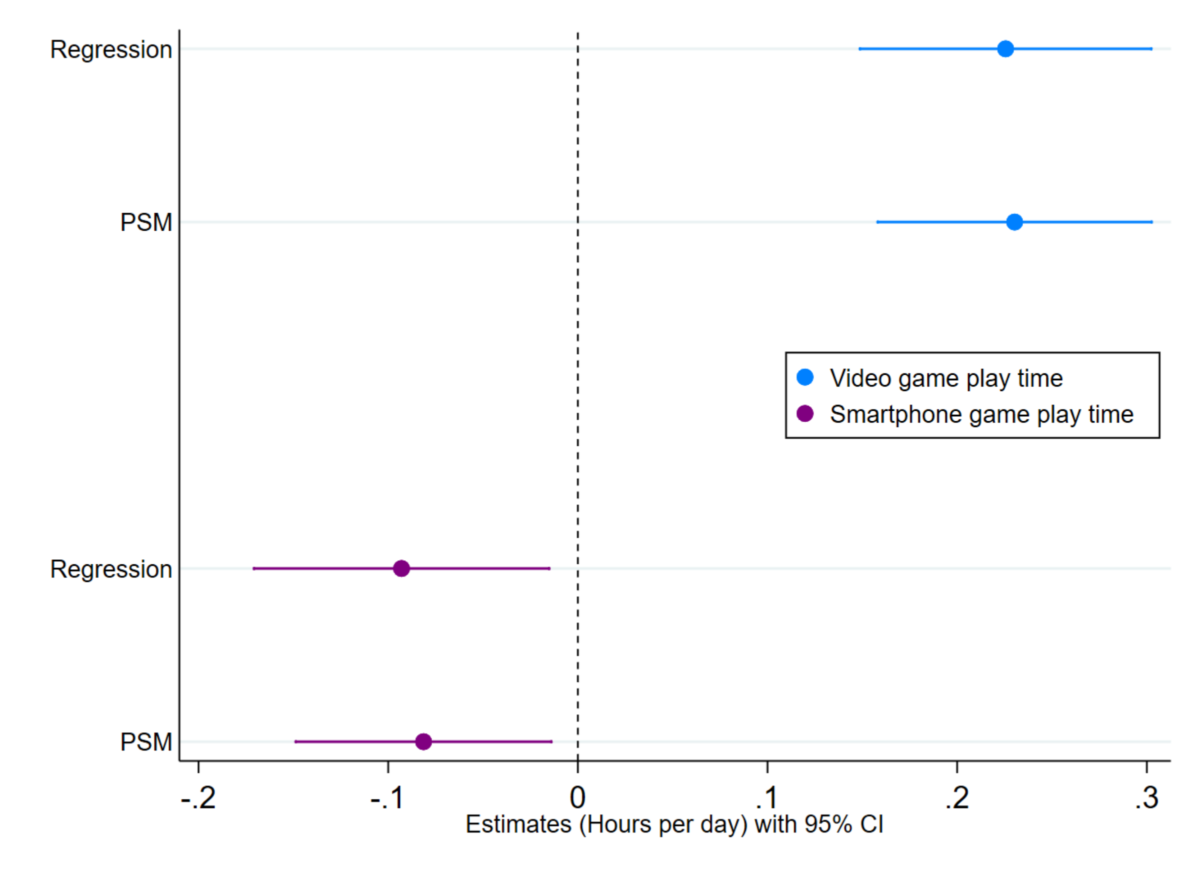


**Supplementary Figure 2: Causal effect of winning game console lotteries on gaming time (N=6,911).**

Notes. CI, confidence interval; PSM, Propensity Score Matching. The analysis sample was limited to those who joined game console lotteries. The point estimates (mean values) and the 95 percent confidence intervals are shown. The regression estimates used Equation (1) and standard errors were clustered by prefectures. Abadie-Imbens robust standard errors were used for PSM.

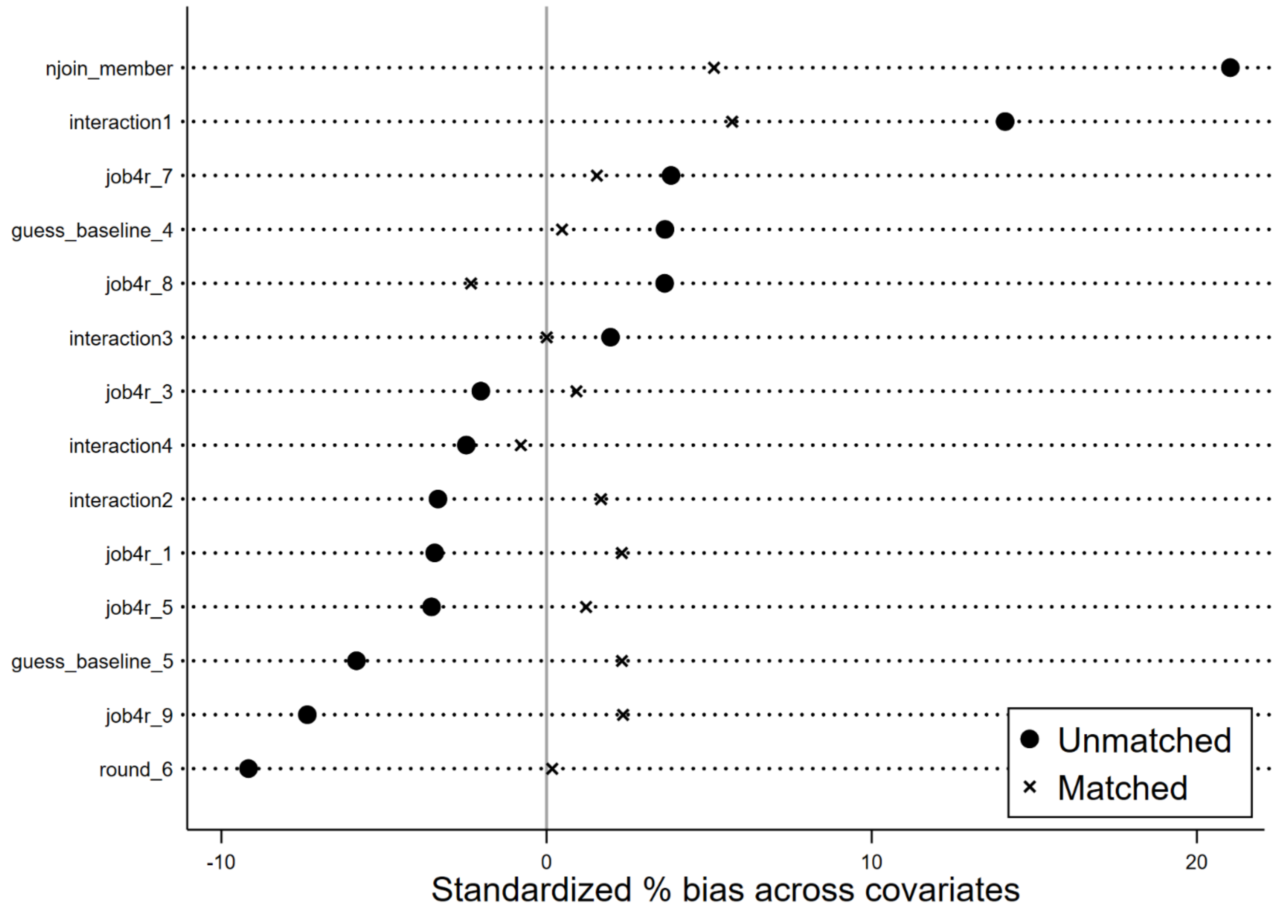


**Supplementary Figure 3: Balance check before and after propensity score matching for estimations.**

Notes. njoin_member: The number of times a household joined PlayStation5 lotteries.
round_6: A dummy variable indicating survey round 6.
guess_baseline_4: A dummy variable indicating a casual gamer.
guess_baseline_5: A dummy variable indicating a non-gamer.
job4r_1: A dummy variable indicating that he/she worked for the engineering and construction industry.
job4r_3: A dummy variable indicating that he/she worked for the manufacturing industry.
job4r_5: A dummy variable indicating that he/she worked for distributors and retailers.
job4r_7: A dummy variable indicating that he/she worked for public works.
job4r_8: A dummy variable indicating that he/she worked for IT industries.
job4r_9: A dummy variable indicating that he/she worked for Banks and financial services.
interaction1 = njoin_member * njoin_member
interaction2 = job4r_7 * round_6
interaction3 = job4r_5 * guess_baseline_5
interaction4 = job4r_9 * round_6

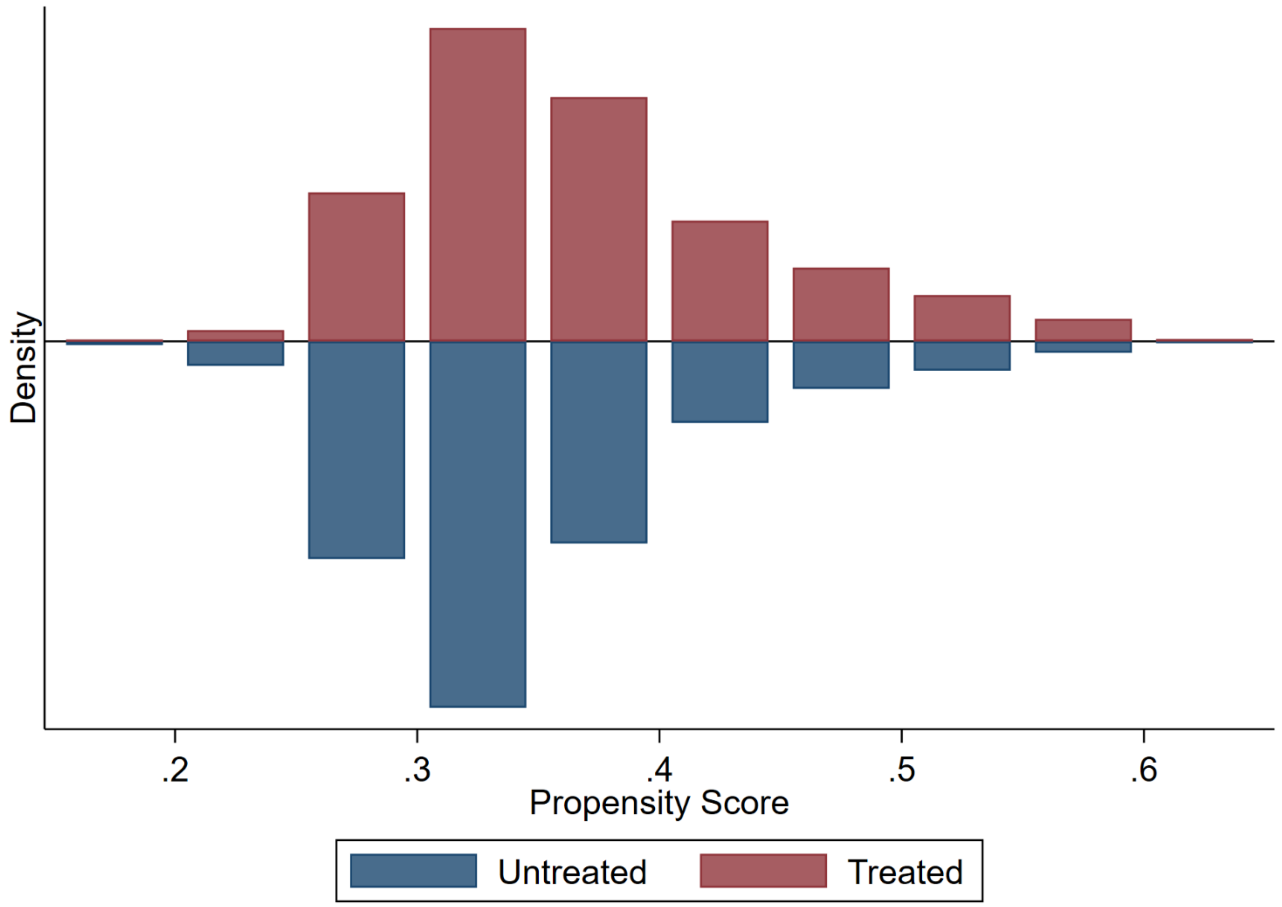


**Supplementary Figure 4: Common support check for propensity score matching estimations.**

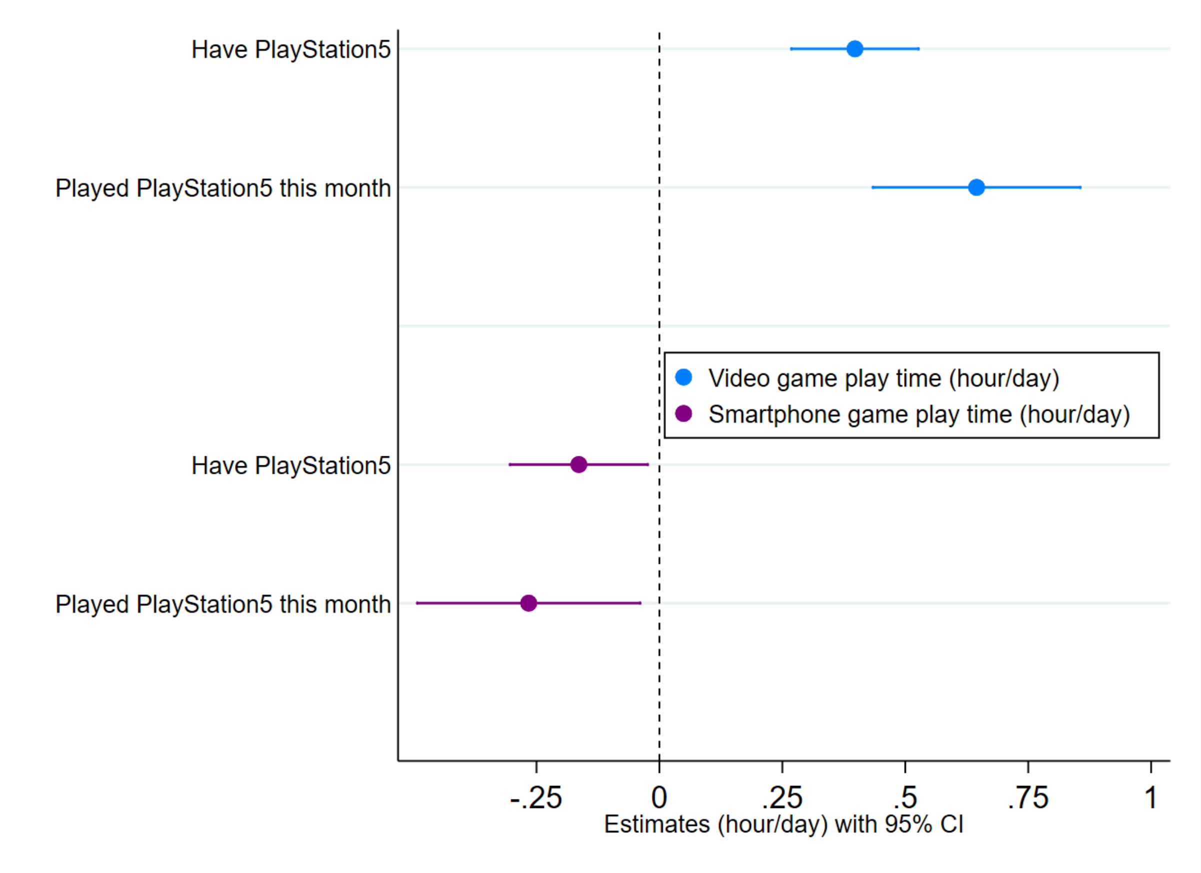


**Supplementary Figure 5: Causal effect of ownership of PS5 on gaming time estimated by instrumental variable method (N=6,911).**

Notes. CI, confidence intervals; PS5, PlayStation5. The analysis sample was limited to those who joined game console lotteries. The point estimates (mean values) and the 95 percent confidence intervals are shown. Standard errors were clustered by prefectures.

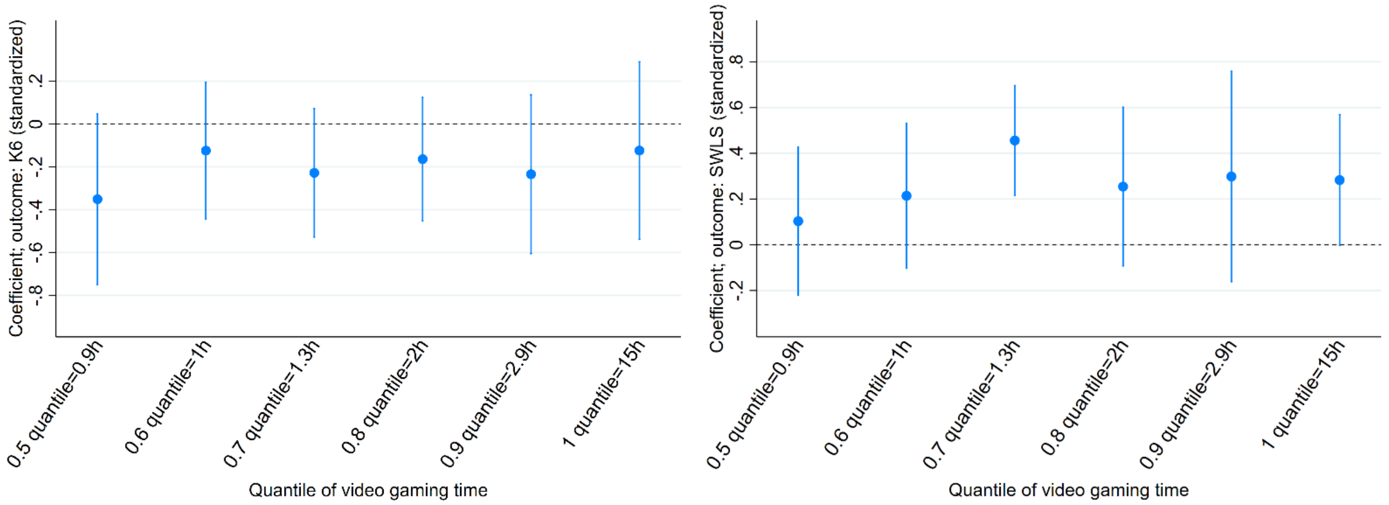


(a) Video gameplay's effect on K6 (b) Video gameplay's effect on SWLS

**Supplementary Figure 6: Subgroup analysis of treatment effect using instrumental variable method.**

Notes. K6, Kessler psychological distress scale; SWLS, The Satisfaction with Life Scale. Treatment effects for subgroups were illustrated, taking β coefficients as the y-axis and the quantile of exposure variables as the x-axis. A lower K6 means having less psychological distress, while a higher SWLS means greater life satisfaction. Subgroup instrumental variable regression bandwidths are 60 percent. Each figure shows estimates based on six subgroups: (i) 0.4 to 0.5 quantile (0.5 to 0.9 hours; n=793), (ii) 0.5 to 0.6 quantile (0.9 to 1.0 hours; n=862), (iii) 0.6 to 0.7 quantile (1.0 to 1.3 hours; n=479), (iv) 0.7 to 0.8 quantile (1.4 to 2 hours; n=878), (v) 0.8 to 0.9 quantile (2.1 to 2.9 hours; n=487), and (vi) 0.9 to 1.0 quantile (2.9 to 15 hours; n=693). The exposure variable was playing PS5 last month. All subgroup estimates had no weak instrument problem (Kleibergen-Paap F statistics are all above 100 for each (i)-(vi). The threshold for detecting a weak instrument is <10.). Regression standard errors were clustered by prefectures. The point estimates (mean values) and the 95 percent confidence intervals are shown.

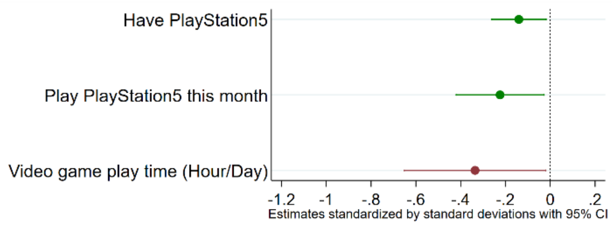


(a) Psychological distress (K6)

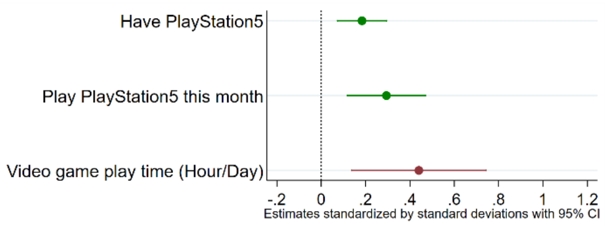


(b) Life satisfaction (SWLS)

**Supplementary Figure 7: Sensitivity Check: Causal effects of video game engagement on mental well-being in post-COVID Japan (N=4,271).**

Notes. CI, confidence intervals. The causal effect of video game engagement on mental well-being is estimated by the IV method. The effects on psychological distress (a) and on life satisfaction (b). The analysis sample comprises rounds 7 and 8 (December 2022 and March 2023), limited to participants who joined game console lotteries. The point estimates (mean values) and the 95% CIs are shown. Standard errors are clustered by prefectures. A lower K6 score indicates less psychological distress, whereas a higher SWLS score indicates greater life satisfaction. The estimates are standardized by the s.d and also available in Supplementary Table 12. The estimates for possession of video game consoles are preferred as they are more likely to adhere to the exclusion restriction requirement, a nontestable assumption of the IV method. The estimates based on console usage and play duration are more prone to violating this assumption.

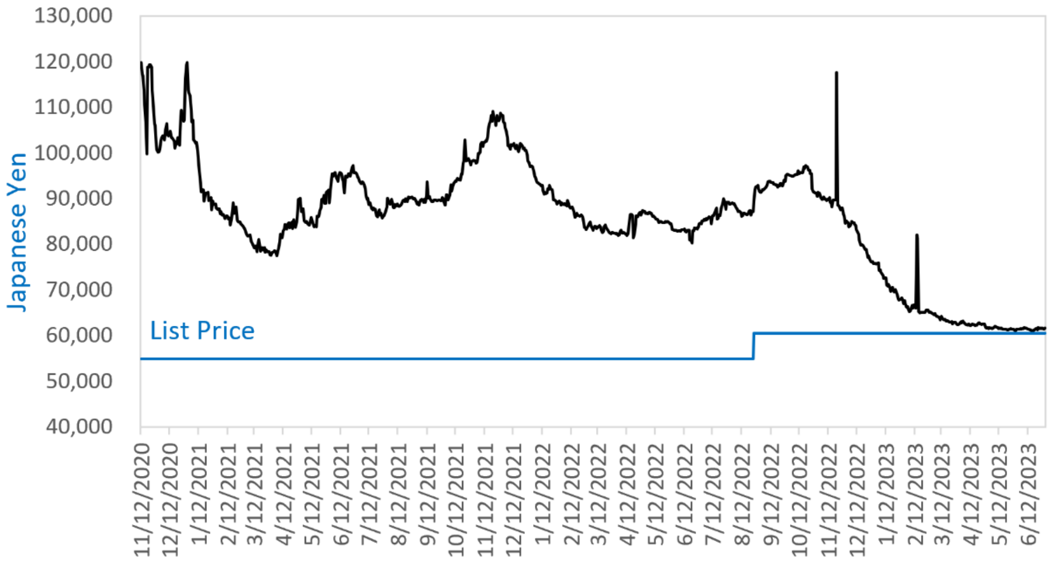


**Supplementary Figure 8: Price history of PS5 (Japanese Yen).**

Notes. PS5, PlayStation5. The average prices are displayed. Source: https://kakaku.com/

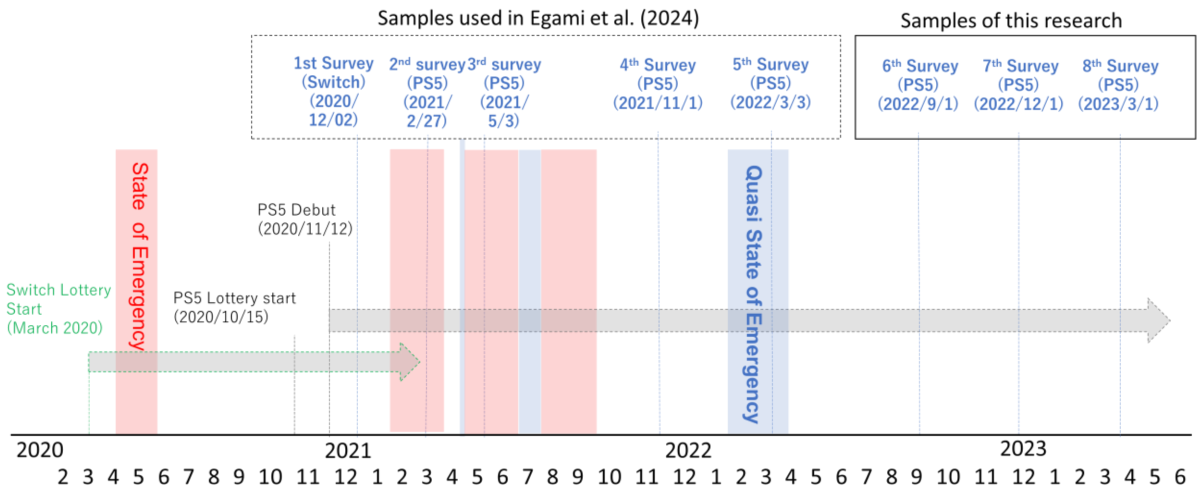


**Supplementary Figure 9: Survey schedule and COVID-19 situation.**
Notes. PS5, PlayStation5. Switch, Nintendo Switch.

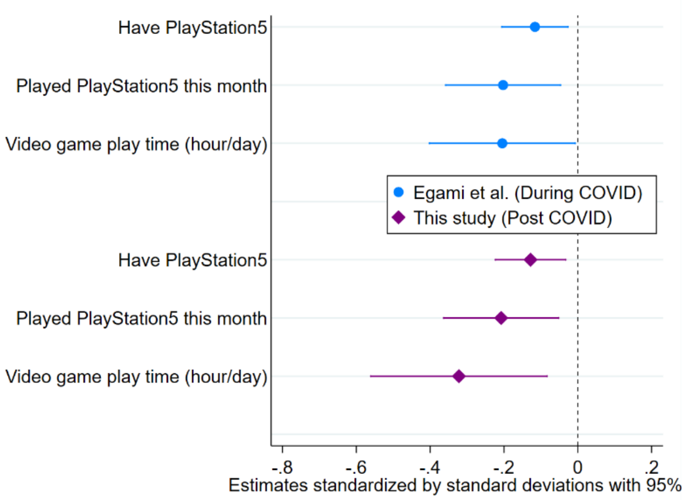


(a) Psychological distress (K6)

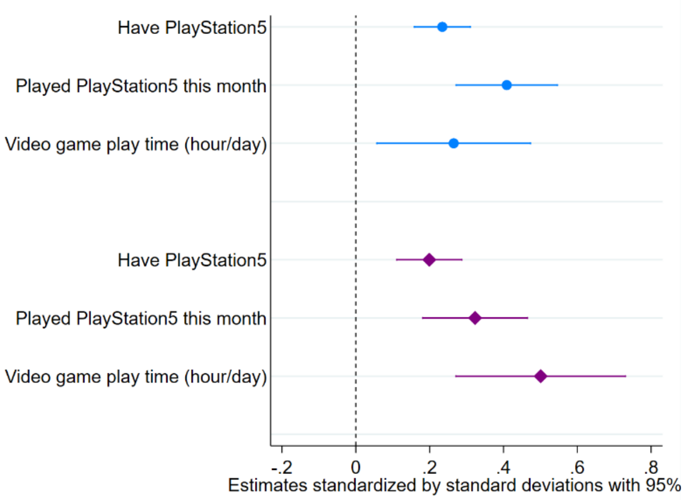


(b) Life satisfaction (SWLS)

**Supplementary Figure 10: Comparison between effects of video game engagement on mental well-being during the COVID-19 and post-COVID-19 period (N=6,911).**

Notes. CI, confidence intervals; PS5, PlayStation5. K6, Kessler psychological distress scale; SWLS, The Satisfaction with Life Scale. The effects on psychological distress (a) and on life satisfaction (b). The causal effect of video game engagement on mental well-being is estimated via the IV method. The point estimates (mean values) and the 95% CIs from Egami et al. and this study are shown. Exposure variables were PS5 ownership, playing PS5 last month, and gameplay duration. Each estimate used winning PlayStation5 lottery as its instrument. The estimates are standardized by the standard deviations. Standard errors are clustered by prefectures. The analysis sample is limited to those who joined game console lotteries. The estimates are also presented in Supplementary Table 9. The estimates for possession of video game consoles are preferred as they are more likely to adhere to the exclusion restriction requirement, a nontestable assumption of the IV method. The estimates based on console usage and play duration are more prone to violating this assumption.

### References.
